# Multisystemic cellular tropism of SARS-CoV-2 in autopsies of COVID-19 patients

**DOI:** 10.1101/2021.06.03.21258241

**Authors:** Dickson W.L. Wong, Barbara M. Klinkhammer, Sonja Djudjaj, Sophia Villwock, M. Cherelle Timm, Eva M. Buhl, Sophie Wucherpfennig, Claudio Cacchi, Till Braunschweig, Ruth Knüchel-Clarke, Danny Jonigk, Christopher Werlein, Roman D. Bülow, Edgar Dahl, Saskia von Stillfried, Peter Boor

## Abstract

**Background:** Multiorgan tropism of SARS-CoV-2 has previously been shown for several major organs.

**Methods:** We have comprehensively analyzed 25 different formalin-fixed paraffin-embedded (FFPE) tissues/organs from autopsies of fatal COVID-19 cases (n=8), using detailed histopathological assessment, detection of SARS-CoV-2 RNA using polymerase chain reaction and RNA *in situ* hybridization, viral protein using immunohistochemistry, and virus particles using transmission electron microscopy. Finally, we confirmed these findings in an independent external autopsy cohort (n=9).

**Findings:** SARS-CoV-2 RNA was mainly localized in epithelial cells, endothelial and mesenchymal cells across all organs. Next to lung, trachea, kidney, heart, or liver, viral RNA was also found in tonsils, salivary glands, oropharynx, thyroid, adrenal gland, testicles, prostate, ovaries, small bowel, lymph nodes, skin and skeletal muscle. Viral RNA was predominantly found in cells expressing ACE2, TMPRSS2, or both. The SARS-CoV-2 replicating RNA was also detected in these organs. Immunohistochemistry and electron microscopy were not suitable for reliable and specific SARS-CoV-2 detection in autopsies. The findings were validated using *in situ* hybridization on external COVID-19 autopsy samples. Finally, apart from the lung, correlation of virus detection and histopathological assessment did not reveal any specific alterations that could be attributed to SARS-CoV-2.

**Interpretation:** SARS-CoV-2 could be observed in virtually all organs, colocalizing with ACE2 and TMPRSS2 mainly in epithelial but also in mesenchymal and endothelial cells, and viral replication was found across all organ systems. Apart from the respiratory tract, no specific (histo-)morphologic alterations could be assigned to the SARS-CoV-2 infection.

**Research in context:** *Evidence before this study:* SARS-CoV-2 has been shown to infect the respiratory tract and affect several other major organs. However, on a cellular level, the localization of SARS-CoV-2 and its targets ACE2 and TMPRSS2 have not been described comprehensively.

*Added value of this study:* We have analyzed tissue SARS-CoV-2 RNA using RT-PCR and visualized its localization together with ACE2 and TMPRSS2 using *in situ* hybridization (ISH) in 25 different autopsy tissues. SARS-CoV-2 sense and antisense RNA were detected in 16 tissues/organs, mainly in epithelial cells and, to a lesser extent, in endothelial or stromal cells. Detection of viral protein using immunohistochemistry or viral particles using transmission electron microscopy did not yield specific results. Interestingly, apart from the respiratory tract and specifically the lungs, we have not found a specific pathology that would be associated with extrapulmonary viral spread.

*Implications of all the available evidence:* We provide a recommendation on using these methods in autopsy diagnostics for SARS-CoV-2. Our data extend the current hypothesis of severe COVID-19 being multisystemic diseases. Our data also provide clear evidence of infection and replication of SARS-CoV-2 in the endothelial cell across all organs, extending the hypothesis on the (micro)vascular involvement in COVID-19.

## Introduction

Since late 2019, the severe acute respiratory syndrome coronavirus 2 (SARS-CoV-2) causing the Coronavirus Disease 2019 (COVID-19) has caused a pandemic with major impacts on virtually all aspects of life. By May 2021, the number of global deaths attributed to COVID-19 has increased to more than 3 million, with more than 160 million infected individuals in 223 countries (WHO; https://covid19.who.int/). Despite extensive research, understanding of the organic and cellular manifestations of COVID-19 remains incomplete. Next to the typical clinical symptoms of a respiratory infection ^1^, including fever, dyspnea, and dry cough, other symptoms indicate the involvement of other organ systems, e.g., diarrhea, vomiting, or anosmia. In severe cases, patients develop acute respiratory distress syndrome (ARDS), pneumonia but also systemic inflammation, heart diseases (e.g. elevated cardiac markers, acute heart failure) ^2^, and acute kidney injury (AKI). These data suggest that COVID-19 can be viewed as a multisystemic disease.

Post-mortem investigation, i.e., autopsy, provides a comprehensive insight into the pathophysiology of novel diseases, such as COVID-19, allowing investigation of the multisystemic viral spread and effects on a tissue and cellular level ^3^. Recent studies using *in situ* hybridization (ISH) have found SARS-CoV-2 tropism and replication in airways, i.e., trachea, lung, bronchi, or submucosal glands, within pneumocytes, alveolar and pulmonary lymph node macrophages, endothelium, and respiratory epithelium ^4-11^. In extra-respiratory organs, ISH demonstrated direct infection of vascular endothelium in the kidney (renal proximal and distal tubular epithelial cells) ^9,12-14^, heart ^10^, liver ^9^, brain stem and leptomeninges ^9^ and placenta (syncytiotrophoblast and cytotrophoblast) ^15^. However, the tissue and cellular association of SARS-CoV-2 genomic sense RNA and antisense RNA (indicating replication), with the receptor and protease required for viral entry into the cells, i.e. angiotensin-converting enzyme 2 (ACE2) and transmembrane protease serine subtype 2 (TMPRSS2), across the various tissues were not comprehensively described ^16^. Here, we used a multitude of different methods to analyze the distribution of SARS-CoV-2 virus, ACE2, and TMPRSS2 in 25 different organs, using formalin-fixed paraffin-embedded (FFPE) autopsy specimens from deceased COVID-19 patients.

## Methods

### Ethical issues

The study was approved by the local ethics committee (EK 304/20, EK 119/20, EK 092/20 and 9621_BO_K_2021) and was carried out in accordance with the declaration of Helsinki for medical research involving human subjects. For all autopsies, legal authorization was obtained from the next of kin of the deceased person.

### Histology samples & cohort description

We used tissue collected from eight autopsies (Table 1). At the time of death, patients were 68±9·55 years old and predominantly male (M:F = 5:3). All patients tested positive for SARS-CoV-2 by RT-PCR during their clinical course. After death, corpses were cooled at 4°C from 4 h after death until autopsy. The post-mortem interval (i.e., time from death to autopsy) ranged from 20 to 48 h (median, 35·5 h). Autopsies were performed in a dedicated room equipped with a separate ventilation system. All autopsies were performed in two steps, with internal organs removed first and then fixed in 4% buffered formalin and without brain autopsy to avoid aerosol formation. Organs were fixed for one week before further processing. Then, all organs from the thorax and abdomen were examined macroscopically and microscopically (tongue, oropharynx, tonsil, salivary gland, thyroid, proximal and distal trachea, paraaortal/ cervical/ hilar/ abdominal lymph nodes, each lung lobe (one central and one peripheral sample each), heart (anterior, lateral and posterior left ventricular wall, septum, left ventricular papillary muscles, and right ventricle), esophagus, stomach, small intestine, colon, left and right liver lobes, pancreas, spleen, left and right kidney, left and right adrenal gland, uterus (f)/ prostate (m), ovary (f)/ testis (m), muscle, and skin. Personal protective equipment included N95 masks, waterproof protective suits, goggles, waterproof aprons, and multiple layers of gloves. All garments were changed before and after autopsies were performed. The detailed protocol of the autopsy procedure is available at https://www.pathologie-dgp.de/media/Dgp/aktuelles/Anl._3_Obduktion_COVID-19_150420201_.pdf (in german). We have also used an independent validation cohort of nine cases described in detail in the supplementary methods.

**Table 1.** Patient characteristics of our COVID-19 study cohort. BMI = Body-mass index; d = day; y = year

| Patient | Sex | Age range (y) | BMI | Smoker | First symptoms to death (d) | Hospital admission to death (d) | Death to autopsy (d) | Duration of ventilation therapy (d) |
| --- | --- | --- | --- | --- | --- | --- | --- | --- |
| 1 | M | 76-80 | Moderately obese | Unknown | 7 | 2 | 1 | 0 |
| 2 | F | 71-75 | Overweight | Unknown | 12 | 7 | 1 | 7 |
| 3 | M | 81-85 | Overweight | Personal history of tobacco abuse | 12 | 11 | 1 | 7 |
| 4 | M | 56-60 | Normal | Unknown | 27 | 24 | 1 | 24 |
| 5 | M | 66-70 | Overweight | Unknown | 35 | 29 | 1 | 29 |
| 6 | F | 71-75 | Overweight | Unknown | 38 | 33 | 2 | 32 |
| 7 | M | 55-60 | Normal | Unknown | 52 | 46 | 2 | 44 |
| 8 | F | 61-65 | Normal | Unknown | 64 | 60 | 2 | 57 |

### Histopathological examination

For microscopic examination, all formalin-fixed specimens were dehydrated and paraffin-embedded (formalin fixed and paraffin embedded -FFPE), cut into 4 µm thin slices, and stained on an autostainer according to standard protocols with hematoxylin-eosin (HE) stain. All slides were digitized using the Aperio AT2 brightfield scanner (Leica Biosystems, Nussloch, Germany) with a 40x objective generating Whole Slides Images (WSI).

### Tissue microarray (TMA) block preparation

For FISH analysis, tissue microarrays were created from 25 organs (oropharynx, tonsil, salivary gland, thyroid, distal trachea, perihilar/ mesenteric lymph node, lung (left and right, two central and one peripheral), heart (anterior, left ventricle wall), esophagus, stomach, large/small intestine, liver, pancreas, spleen, kidney, adrenal gland, urinary bladder, uterus/prostate, ovary/testis, bone marrow, muscle, and skin; Supplementary Table 1). Punches for tissue microarrays (3 mm diameter) were taken under HE-morphologic control to obtain areas with the least autolytic changes or most representative areas for specific pathologic changes (e.g., hyaline membranes, squamous metaplasia in the lung, neoplastic cells in malignant disease). TMAs were then sectioned with a microtome sequentially for various stains, including hematoxylin-eosin (HE) and *in situ* hybridization, as mentioned below.

### SARS-CoV-2 RNA detection with Fluorescence *in situ* hybridization (FISH)

The fresh-cut, 1-μm-thick TMA sections were deparaffinized in xylene and then dehydrated with 100% ethanol. FISH was performed on the sections with the RNAscope® Multiplex Fluorescent Reagent Kit v2 assay (Advanced Cell Diagnostics, Inc., Hayward, California), based on the assay principle as described ^17^. Briefly, we incubated the tissue sections with H_2_O_2_ and performed a heat-induced target retrieval step followed by protease incubation with the reagents provided. RNA sequences of SARS-CoV-2 S gene, SARS-CoV-2 antisense of S gene, ACE2, and TMPRSS2 were hybridized using RNAscope® probe -V-nCoV2019-S (#848561-C1), -V-nCoV2019-S-sense (#845701-C1), -Hs-ACE2-C2 (#848151-C2) and -Hs-TMPRSS2-C2 (#470341-C2), respectively. Positive (C1: *POLR2A* gene of *Homo sapiens*; C2: *PPIB* gene of *Homo sapiens*) and negative probes (*dap* gene of *Bacillus subtilis*) were also applied in each experiment. After the amplifier steps, according to the manual, Opal™ 570 and 650 fluorophores (PerkinElmer Life and Analytical Sciences, Boston, MA) were applied to the tissues incubated with C1 and C2 probes, respectively. Finally, nuclei were labeled with DAPI, and the slides were mounted with ProLong™ Gold antifade reagent (Invitrogen, Waltham, MA). Sections were analyzed with Zeiss Axio Imager 2 and image analysis software (ZEN 3·0 blue edition).

### Electron Microscopy (EM)

Samples prepared for transmission electron microscopy were obtained after 7-10 days of formalin fixation and additional fixation in 3% glutaraldehyde for 24 hours. Samples were post-fixed in 1% OsO_4_ (Roth, Karlsruhe, Germany) and dehydrated in ascending ethanol series (30, 50, 70, 90, and 100%). Dehydrated samples were incubated in propylene oxide (Serva, Heidelberg, Germany) and embedded in Epon resin (Serva, Heidelberg, Germany). Ultrathin sections were contrast-enhanced by staining with 0.5% uranyl acetate and 1% lead citrate (both EMS, Munich, Germany) and viewed on a transmission electron microscope (Zeiss Leo 906, Oberkochen, Germany).

### Immunohistochemical (IHC) staining of SARS-CoV-2 spike glycoprotein in FFPE autopsy lung

We selected autopsy lung samples from a cohort of 3 patient groups, which were diagnosed as ARDS, COVID-19, and influenza A virus subtype H1N1 infection (Table 2). Briefly, 4 μm thin FFPE sections were prepared and deparaffinized in xylene, followed by rehydration with a concentration gradient of ethanol. Tissue sections were then subjected to heat-induced epitope retrieval with Tris-EDTA buffer (pH 9) and quenched with 3% H_2_O_2_. The slides were then incubated with primary antibody against SARS spike glycoprotein (1:1000; Abcam, Cambridge, UK; Ab272420) in 1% BSA/PBS solution. The remaining IHC steps were performed as previously described ^18^, and the staining was developed with ImmPACT® VIP Substrate (Vector Labs, Burlingame, CA, US).

**Table 2.** List of lung autopsy cohort for testing IHC staining of SARS-CoV-2 spike glycoprotein.

| Diagnosis | Sample ID | Autopsy date | Diffuse alveolar damage (DAD) phase | Spike protein stain |
| --- | --- | --- | --- | --- |
| ARDS<br>(n=6) | IHC1 | June 2010 | Exudative/ hyaline membranes | Diffuse |
|  | IHC2 | April 2011 | Fibrotic phase | Diffuse |
|  | IHC3 | March 2014 | Fibrotic phase | Diffuse |
|  | IHC4 | January 2018 | Exudative/ hyaline membranes | Diffuse |
|  | IHC5 | February 2019 | Fibrotic phase | Few positive cells |
|  | IHC6 | July 2019 | Fibrotic phase | Few positive cells |
| Influenza A<br>(H1N1)<br>(n=4) | IHC7 | November 2009 | Proliferative/ organizing phase | Diffuse |
|  | IHC8 | March 2010 | Proliferative/ organizing phase | Diffuse |
|  | IHC9 | February 2018 | Proliferative/ organizing phase | Diffuse |
|  | IHC10 | March 2018 | Proliferative/ organizing phase | Few positive cells |
| COVID-19<br>(n=6) | Patient 1 | April 2020 | Proliferative/ organizing phase | Diffuse |
|  | Patient 4 | April 2020 | Fibrotic phase | Focal |
|  | Patient 5 | April 2020 | Proliferative/ organizing phase | Diffuse |
|  | Patient 6 | May 2020 | Proliferative/ organizing phase | Diffuse |
|  | Patient 7 | May 2020 | Proliferative/ organizing phase | Focal |
|  | Patient 8 | May 2020 | No DAD | Diffuse |
| Normal<br>(n=6) | IHC17 | June 2009 | Normal | Negative |
|  | IHC18 | June 2009 | Normal | Few positive cells |
|  | IHC19 | May 2011 | Normal/ edema | Diffuse |
|  | IHC20 | April 2013 | Normal | Diffuse |
|  | IHC21 | January 2015 | Atelectasis | Diffuse |
|  | IHC22 | June 2015 | Normal | Some positive cells |

### RNA isolation from FFPE specimens and SARS-CoV-2 RNA detection

We extracted RNA from FFPE tissue using a Maxwell® 16 LEV RNA FFPE Purification Kit (Promega GmbH, Walldorf, Germany) on the Maxwell® 16 IVD instrument (Promega GmbH) or with the ReliaPrep™ FFPE Total RNA Miniprep System (Promega GmbH) according to the manufacturer’s instructions. We stored the RNA samples at -80°C until further processing.

Previously we compared two different kits for RT-PCR analysis ^19^. Here, we used the TaqMan(tm) Fast 1-Step Master Mix (Thermo Fisher Scientific GmbH, Dreieich, Germany) for the qualitative detection of the E gene (encoding envelope protein) of severe acute respiratory syndrome coronavirus type 2 (SARS-CoV-2) by a primer (0·4 μM) and probe (0·2 μM) set labeled with fluorescent reporters and quencher dyes. We used TaqMan® Exogenous Internal Positive Control reagents (Thermo Fisher Scientific GmbH, Dreieich, Germany) as internal PCR controls. RT-PCR was performed as previously described ^19,20^. Briefly, we reversely transcribed (50°C for 10 min) and amplified the RNA extracted from FFPE tissues with the reaction mixture at 95°C for 20 s and followed by 45 cycles of 95°C for 3 s and 58°C for 30 s. We used the Amplirun® SARS-CoV-2 RNA control (Bestbion dx GmbH, Cologne, Germany) provided with 13000 viral RNA copies μL^-1^ to calculate the viral RNA copies in the tissue samples. In our SARS-CoV-2 positive tissues, the third quartile of the detectable viral copies is 328 viral copies μL^-1^, which is equivalent to the Ct value of 30·7. In this regard, we defined the cut-off value indicating a high viral load of SARS-CoV-2 in the sample when the Ct value was ≤ 30·7.

## Results

### Study cohort

The main characteristics of the study cohort (5 male/ 3 female; median age 69 [55– 83] years) are given in Table 1. The period between the onset of symptoms and death ranged from 12 to 64 days (median 31 days, Figure 1), thereby including early and late/prolonged disease stages. The time between the onset of symptoms and admission ranged from 1 to 7 days (median 5 days, Figure 1) and from admission to death from 2 to 60 days (median 26·5 days, Figure 1). Six patients had at least one clinically confirmed comorbidity, including hypertension (n=5), diabetes mellitus type 2 (n=3) or type 1 (n=1), coronary artery disease (n=2), malignant disease (n=1), liver cirrhosis (n=1) and dementia (n=1). One patient had a history of chronic kidney disease with kidney transplant and immunosuppression. One patient had a history of tobacco abuse, while no information on current or previous tobacco abuse was available for the remaining 7 patients. None of the patients had tested positive for the respiratory syncytial virus or influenza A and B viruses on admission. Seven patients were treated with mechanical ventilation, of which three patients were treated with additional extracorporeal membrane oxygenation (ECMO) subsequently. One patient refused mechanical ventilation and intensive care treatment. Duration of ECMO treatment ranged from 342 to 1381 hours (median 568 hours). During treatment, five patients had an acute kidney injury and required dialysis. None of the patients had a cerebrovascular disease or chronic obstructive pulmonary disease (COPD).

**Figure 1.**
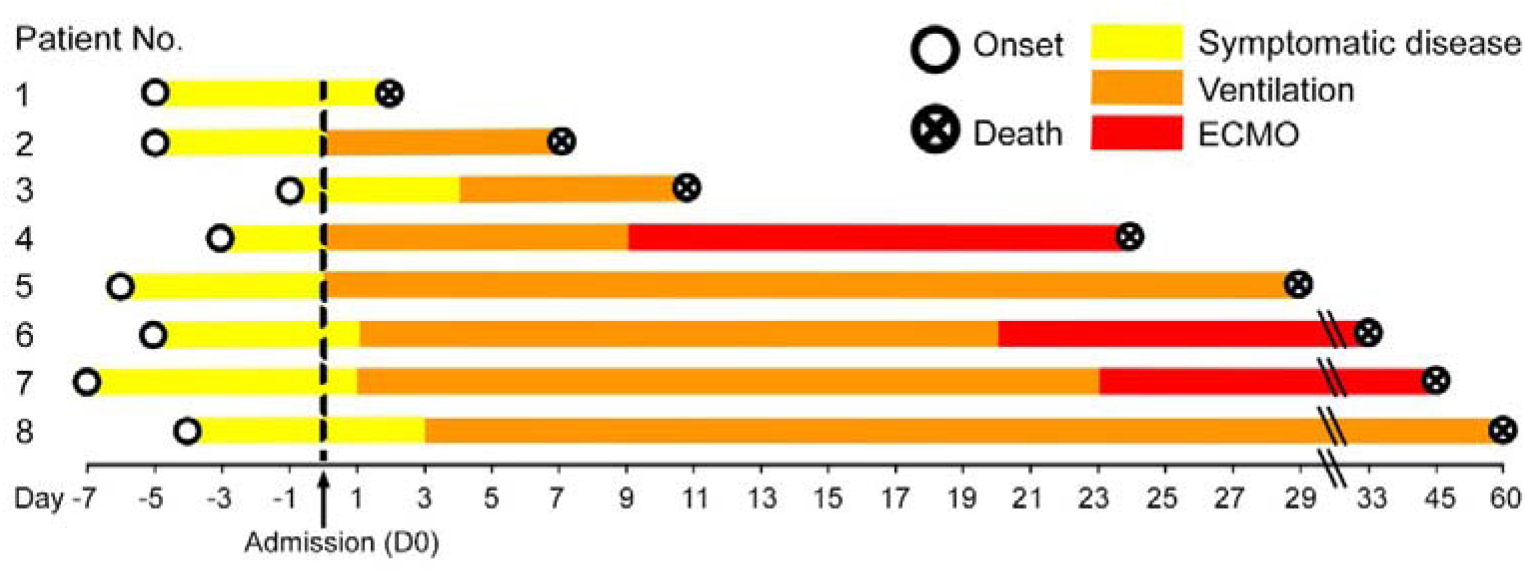
Disease duration of the patient cohort in our study. The disease course of each of the eight COVID-19 patients is shown from disease onset until death. Hospital admission was denoted as day 0, the therapies are color-coded. Patient 1 had the shortest time between the beginning of symptomatic disease and death, i.e. six days, while patient 8 had the longest disease course till death with eight weeks of mechanical ventilation.

### Pathological findings

From the autopsy analyses, the direct cause of death in all patients was diffuse alveolar damage (DAD) with mixed and overlapping patterns of the exudative, proliferative, and organizing phase or fibrotic phase (Supplementary Table 2). All patients had heavier lungs compared to the normal range (median left: 945 g and right: 1191 g, normal: 500-700 g). Intra-alveolar granulocyte infiltration indicating bacterial superinfection was found in the majority of cases (5/8). Additionally, squamous metaplasia (6/8) and multinucleated intra-alveolar giant cells (5/8) were found. Two patients displayed pulmonary artery thrombosis. Some common findings in other organs were ductular cholestasis in the liver (5/8) and cardiomegaly (6/8; median 482 g). All pathological diagnostic data are summarized in Supplementary Table 2.

Pulmonary findings of DAD included hyaline membranes, intra-alveolar edema, proliferative DAD after a disease duration of <21 days in three patients and organizing or fibrotic DAD with bronchiolization and squamous metaplasia after a disease duration of >21 days in five patients (Supplementary Table 2). There was a diffuse distribution of different DAD phases and non-affected tissue, with highly fibrotic areas neighboring next-to-normal appearing regions. Erosive tracheobronchitis was present in two patients. Pulmonary lymph nodes showed enlargement and reactive changes with sinus histiocytosis. Pulmonary metastases from breast or papillary thyroid cancer was found in two patients (Supplementary Figure 2C&D).

No overt pathological findings attributable to Sars-CoV-2 infection could be recognized outside of the lung. In a detailed analysis, two of five patients with a disease duration of >21 days showed signs of borderline myocarditis with >14 CD3 positive lymphocytes/ mm^2^ (patient 4, 50 CD3+ cells/mm^2^ and patient 5, 16 CD3+ cells/ mm^2^). In these patients, cardiac tissue was positive for SARS-CoV-2 RNA (Ct value ≥ 30 in patient 4 and Ct value ≤ 30 in patient 5, Figure 2). In three additional patients with a disease duration of 12-64 days, CD3 positive lymphocytes were increased to ≥10/ mm^2^ (patient 3 and 8, 10 CD3+ cells/mm^2^ and patient 7, 11 CD3+ cells/ mm^2^). In two of these three cases, the cardiac tissue was positive for SARS-CoV-2 RNA (Ct value ≤ 30 in patient 3 and Ct value ≥ 30 in patient 7, Figure 2). In the five cases with increased CD3 lymphocytes, also intravascular lymphocytes were increased, consistent with capillaritis (or endotheliitis). In all cases, we found an increased number of intra-and perivascular CD68 positive macrophages. Myocyte injury was not found in any case.

**Figure 2.**
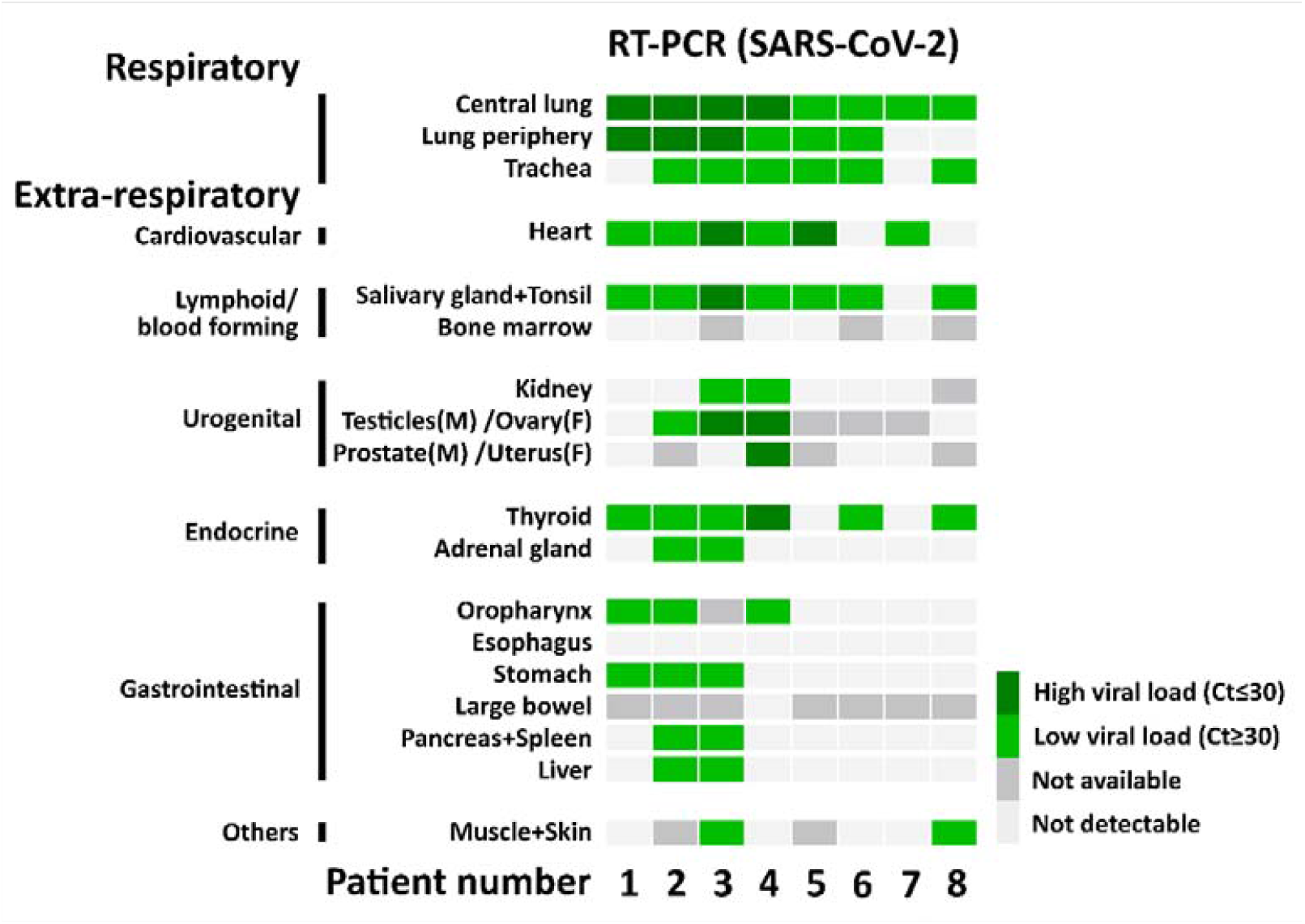
SARS-CoV-2 RNA detection with RT-PCR. The heatmap shows the results of the quantitative PCR detection of SARS-CoV-2 for each patient in all analyzed tissues.

In the haematopoetic system, an increased number of macrophages were visible in the bone marrow of one patient. This patient fulfilled 5 of 8 criteria of the macrophage activation syndrome ^21^. The spleen morphologically seemed depleted of lymphoid cells in all cases.

In the urogenital system, kidney samples from two patients were positive for RT-PCR of SARS-CoV-2 RNA but displayed no specific pathology (Ct value ≥ 30, Patient 3 and 4, Figure 2). In 2/5 males, testicular germ cell aplasia was found. One of these cases was positive for SARS-CoV-2 RNA with high viral load in testicular as well as prostatic tissue (Ct value ≤ 30 in Patient 4, Figure 2), while one another case with high viral load in testicular tissue (Patient 3, Figure 2) did not display morphologic alterations of spermatogenesis. In one case of RT-PCR positive ovarian tissue with low viral load (Ct value ≥ 30, Patient 2, Figure 2), no pathologic findings were noted.

In the endocrine system, we found moderately to highly increased lymphocytic infiltration in the thyroid of 3/8 patients, two of which were also RT-PCR positive for SARS-CoV-2 RNA (Patient 2 and 6, Figure 2). In 2/8 patients, focal inconspicuous chronic inflammatory cells were found, of which one was associated with high viral load (Ct ≤ 30, Patient 4, Figure 2) and one with low viral load (Ct ≥ 30, Patient 8, Figure 2). In one patient, we discovered previously unknown advanced papillary thyroid cancer with pulmonary metastasization (Patient 8), as well as a previously unknown papillary thyroid microcarcinoma of 3 mm diameter in a second patient (Patient 5).

In the adrenal gland, 5/8 cases showed inconspicuous chronic inflammation with perivascular distribution (patient 2, 5-8). One case showed moderate chronic inflammation (Patient 3):Two cases were positive for SARS-CoV-2 RNA with RT-PCR (Ct ≥ 30, Patient 2 and 3, Figure 2).

In the gastrointestinal system, we detected no specific morphologic alterations, despite positive RT-PCR for SARS-CoV-2 RNA in most samples from the salivary gland, oropharynx, stomach, pancreas, and liver in patients with a disease duration < 14 days (Ct ≥ 30, Patients 1-3, Figure 2). The liver in Patient 2 showed micronodular cirrhosis. With an increased disease duration of > 21 days, three of five patients showed ductular cholestasis of the liver (“cholangitis lenta”, Patient 4, 5, and 7, Figure 2), negative for SARS-CoV-2 RNA tested with RT-PCR. Finally, in muscle and skin, we found no specific alterations in two samples positive for SARS-CoV-2 RT-PCR in comparison to those that were negative.

### FISH-based detection of SARS-CoV-2, ACE2, and TMPRSS2

RT-PCR allows fast screening of tissues for SARS-CoV-2 positivity, and thereby effective correlation of pathological findings with viral presence. However, it does not allow cell-specific analyses of localization. For this purpose, we validated a FISH method with positive and negative controls by detecting endogenous human genes *POLR2A* and *PPIB)* and a bacterial gene (*dap* gene of *Bacillus subtilis)*, respectively (Supplementary Figure 1A-E). We also validated the probes against the SARS-CoV-2 S gene and human *ACE2* gene on autopsy lung tissue collected from an influenza A (H1N1) infected patient before the COVID-19 pandemic, showing no signal for SARS-CoV-2 S gene and prominent signal for *ACE2* (Supplementary Figure 1F). FISH was performed to analyze tissue microarrays (TMA), followed by HE staining on consecutive slides for the overlay to allow a detailed morphological correlation (Figure 3-7). We used two probe combinations for the FISH detection, either the SARS-CoV-2 S gene antisense (determining viral genomic RNA) and ACE2 or the SARS-CoV-2-S gene sense strand (indicating replicating virus) and TMPRSS2. Consistent with the RT-PCR results, we identified SARS-CoV-2 positive signals in the lung and respiratory tract (Figure 4, Supplementary Figure 2), heart, lymphoid organs (Figure 5), urogenital tract (Figure 6), endocrine and gastrointestinal organs, muscle and skin (Figure 7). We also observed the positive signal for the replicating SARS-CoV-2 (antisense) in all these organs. The SARS-CoV-2 receptor ACE2 and protease TMPRSS2 RNA were found in all organs except bone marrow and urinary bladder (Figure 3).

**Figure 3.**
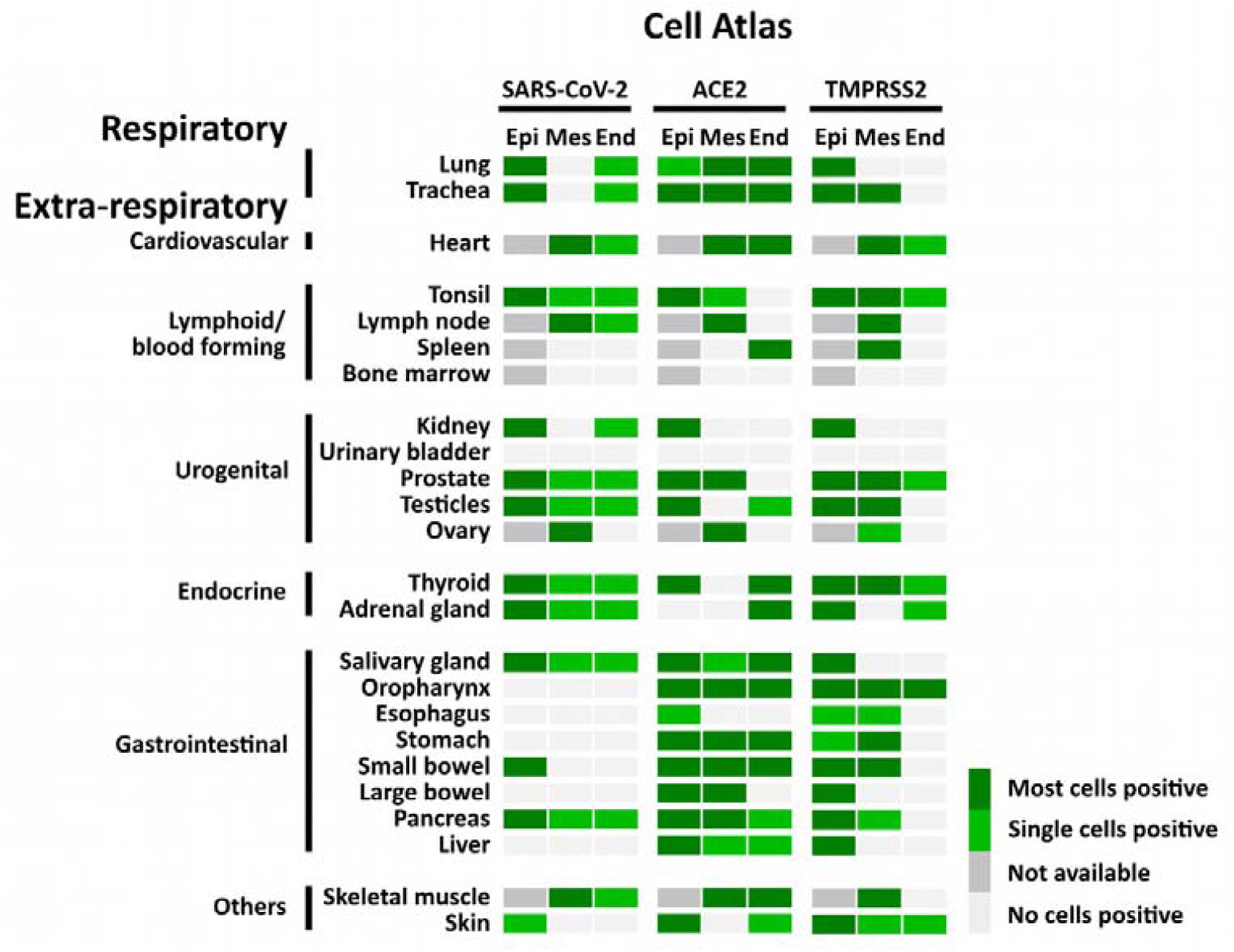
Atlas of cells that were positive for SARS-CoV-2, ACE2, and TMPRSS2 using FISH. The heatmap shows the pattern of SARS-CoV-2, expression of ACE2 and TMPRSS2 in different tissues. Epi = Epithelial cells; Mes = Mesenchymal stromal cells; End = Endothelial cells; Not available = tissue does not contain cell type

**Figure 4.**
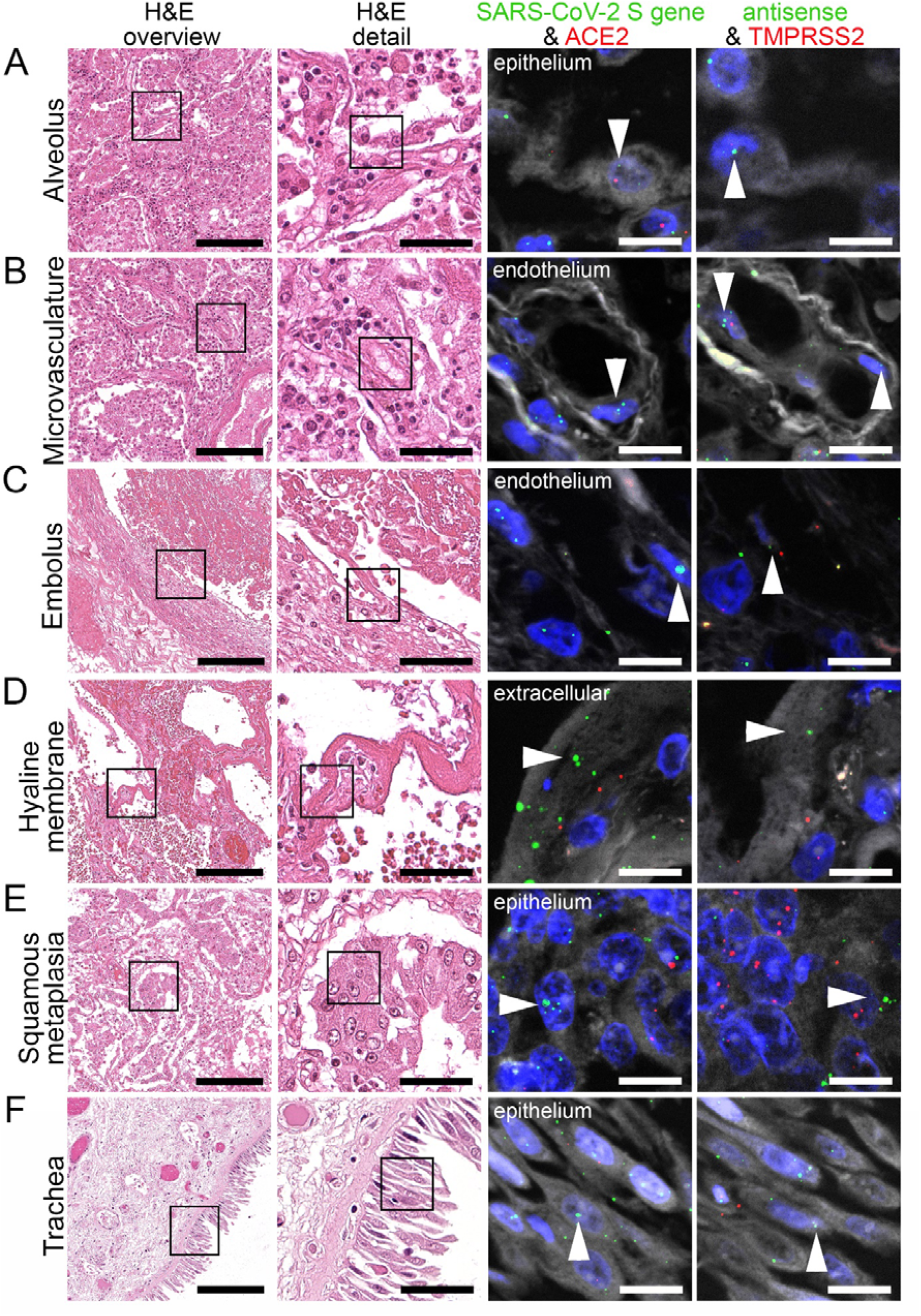
Virus detection in the respiratory system by FISH. HE stained lung tissue and representative image sections showing FISH co-visualization of RNA sequences either of SARS-CoV-2 S gene genomic RNA (green, arrowhead) and ACE2 (red) or SARS-CoV-2 antisense strand RNA as an indicator of replicating virus (green, arrowhead) and TMPRSS2 (red). Morphological details are shown in regions of the alveolus (A, alveolar pneumocytes), endothelium in the alveolar wall (B), endothelial cells adjacent to an embolus (C), hyaline membrane (D), squamous metaplasia (E, metaplastic epithelium), and trachea (F, respiratory epithelium). Scale bars represent 200, 50 and 10 µm, respectively.

**Figure 5.**
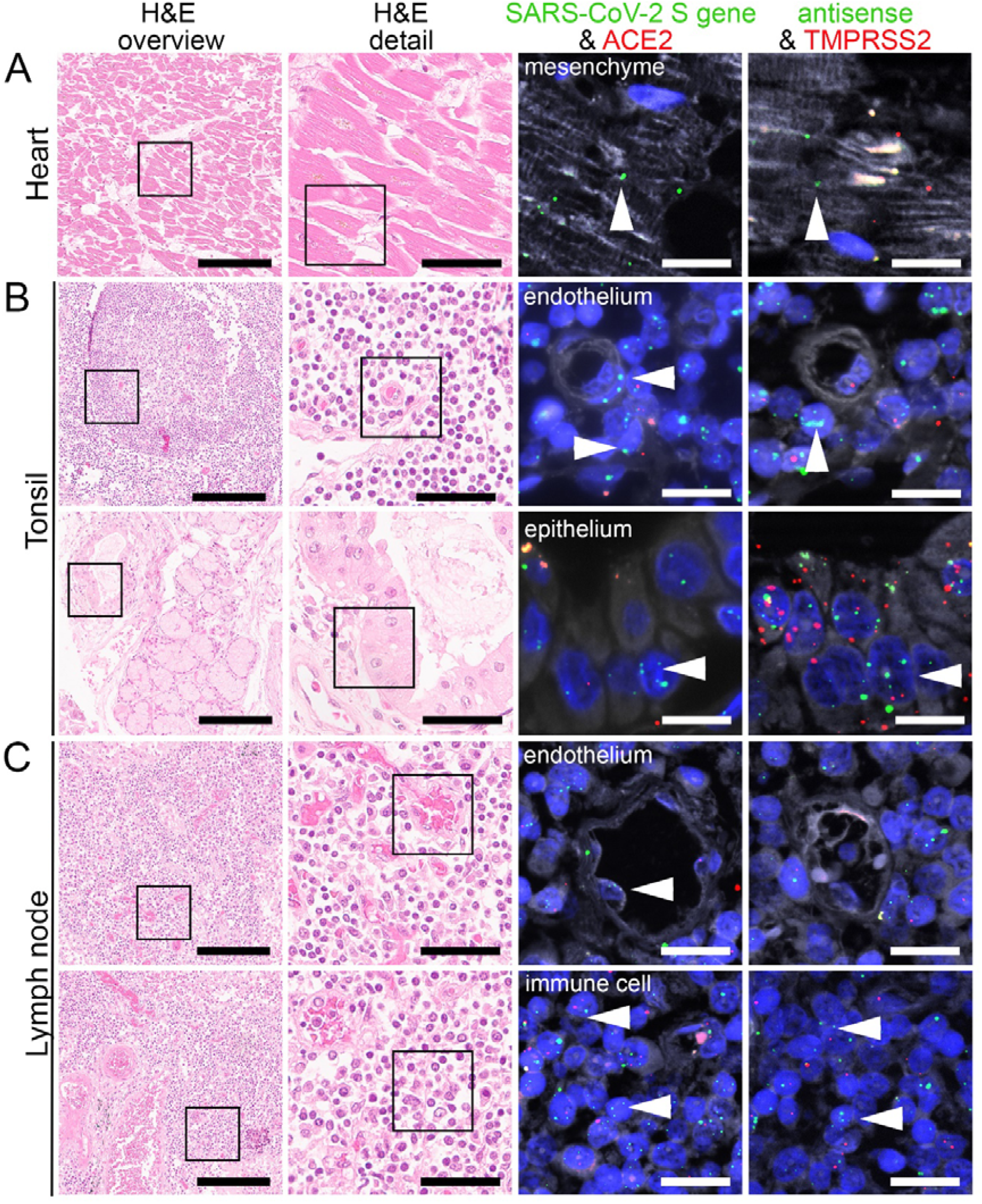
Virus detection in heart and lymphoid organ autopsy tissue by FISH. HE stained tissue and representative image sections showing FISH co-visualization of RNA sequences either of SARS-CoV-2 S gene genomic RNA (green, arrowhead) and ACE2 (red) or SARS-CoV-2 antisense strand RNA indicating replicating virus (green, arrowhead) and TMPRSS2 (red) in the heart (A, cardiomyocyte), tonsil (B, upper panel: capillary with endothelial lining, surrounded by lymphocytes/ immune cells; lower panel: local small salivary gland epithelium) and lymph node (C, upper panel: capillary with endothelial lining; lower panel: lymphocytes/ immune cells). Scale bars represent 200, 50 and 20 µm, respectively.

**Figure 6.**
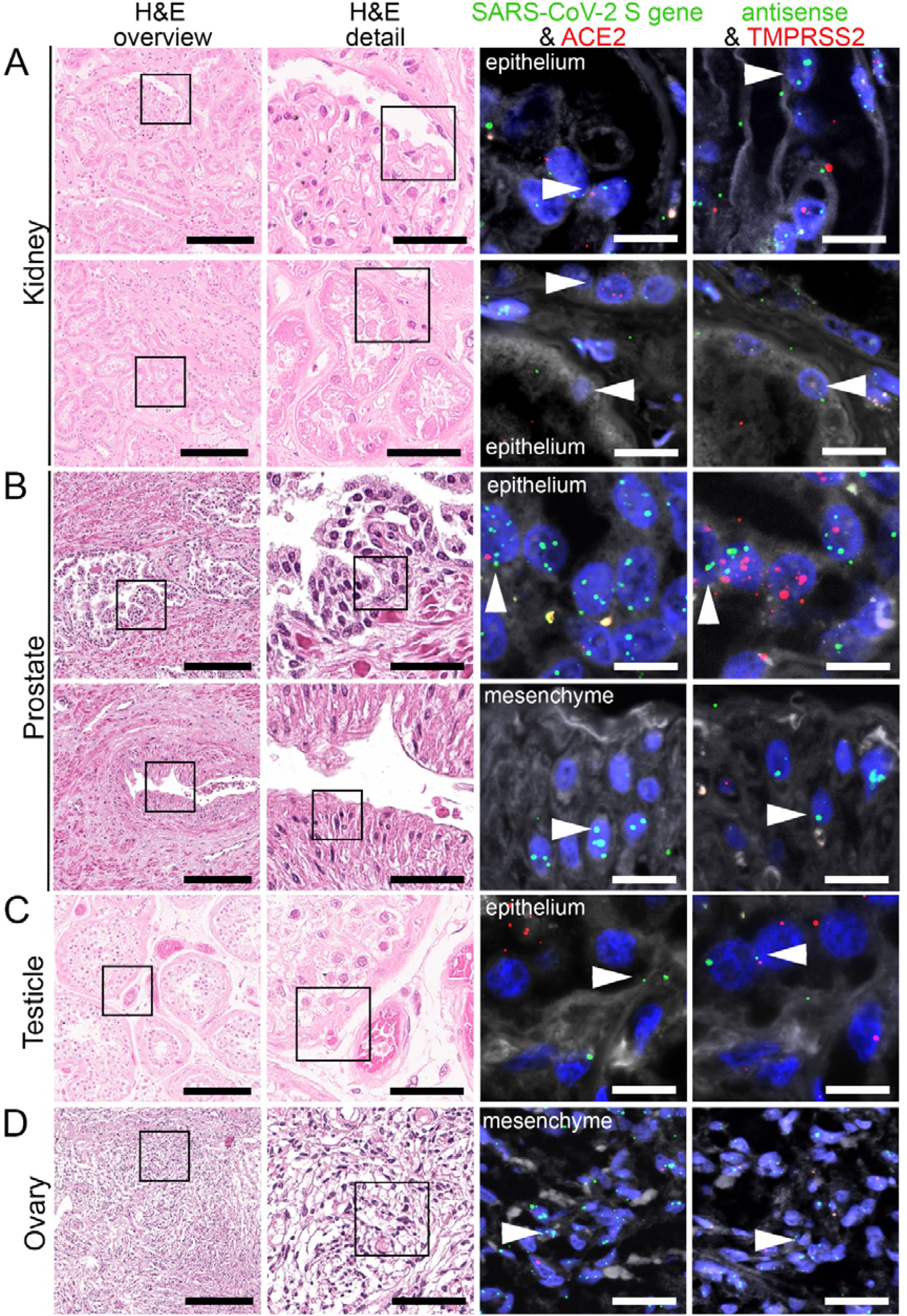
Virus detection in urogenital tract autopsy tissue by FISH. HE stained tissue and representative image sections showing FISH co-visualization of RNA sequences either of SARS-CoV-2 S gene genomic RNA (green, arrowhead) and ACE2 (red) or SARS-CoV-2 antisense strand RNA as an indicator of replicating virus (green, arrowhead) and TMPRSS2 (red) in the kidney (A, upper panel: glomerular visceral epithelial cells/ podocytes; lower panel: tubular epithelial cells), prostate (B, upper panel: glandular epithelial cells; lower panel: vascular smooth muscle cells), testicle (C, germinal epithelium) and ovary (D, mesenchymal stromal cells). Scale bars represent 200, 50 and 20 µm (A, C, D) or 10 µm (B), respectively.

**Figure 7.**
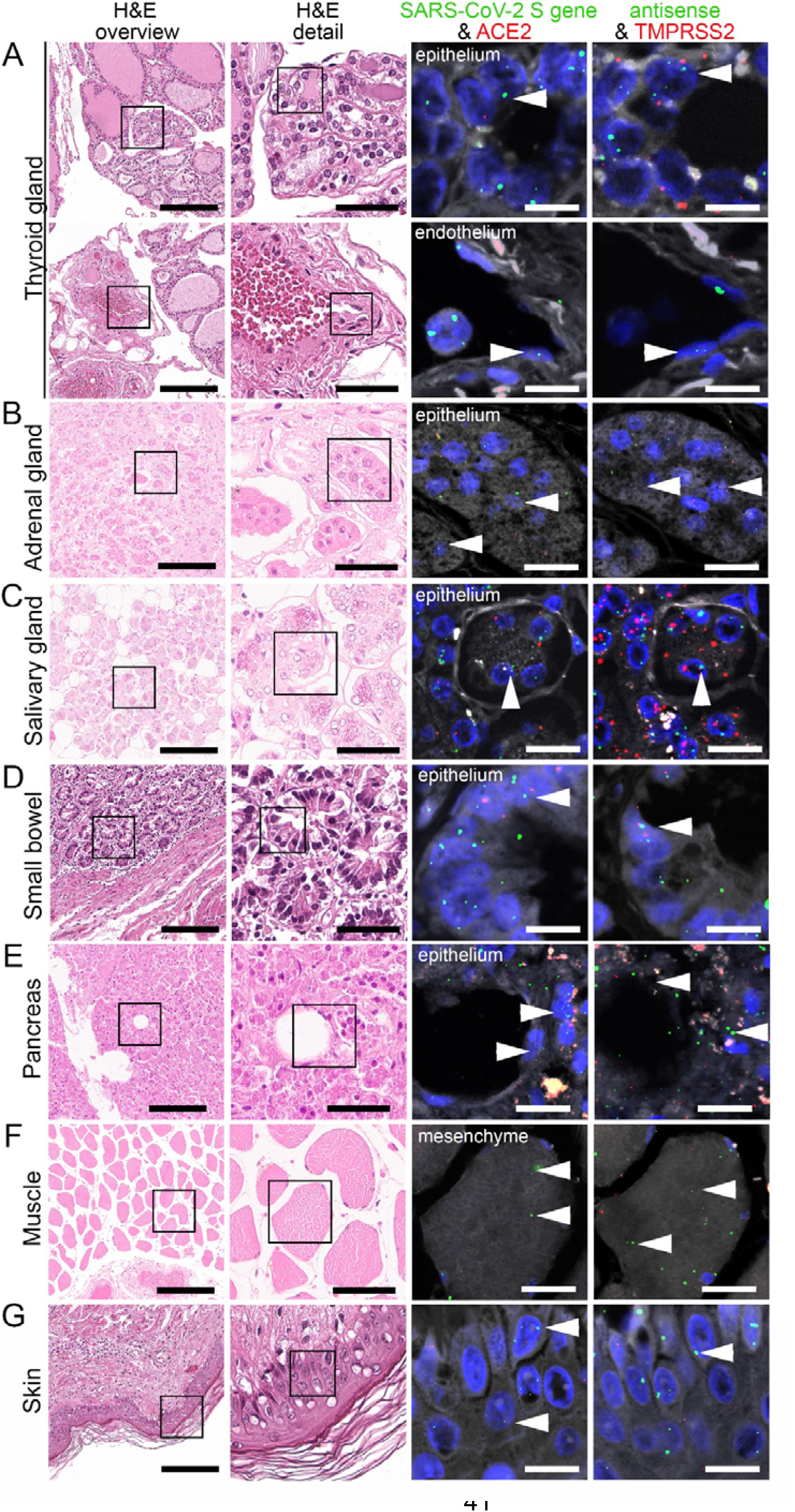
Virus detection in endocrine and gastrointestinal organs and skeletal muscle autopsy tissue by FISH. HE stained tissue and representative image sections showing FISH co-visualization of RNA sequences either of SARS-CoV-2 S gene genomic RNA (green, arrowhead) and ACE2 (red) or SARS-CoV-2 antisense strand RNA indicating replicating virus (green, arrowhead) and TMPRSS2 (red) in the thyroid (A, upper panel: follicular epithelium; lower panel: vascular endothelium), adrenal gland (B, glandular epithelium), salivary gland (C, acini), small bowel (D, crypt epithelium), pancreas (E, acinar epithelium), skeletal muscle (F, skeletal muscle cell) and skin (G). Scale bars represent 200, 50 and 10 µm (A, C, F) or 20 µm (B, D, E), respectively.

### SARS-CoV-2, ACE2, and TMPRSS2 in the respiratory system

We have identified several respiratory cells that were positive for SARS-CoV-2 genomic (S gene; sense) and replicating (S gene: antisense) RNA, ACE2, and TMPRSS2. This included the bronchial and alveolar epithelial cells and cells of the squamous metaplasia in the lung (Figure 4A and 4E, Supplementary Figure 2A). In the microvasculature, SARS-CoV-2 genomic and replicating RNA could be found in the endothelial cells together with ACE2 and TMPRSS2 (Figure 4B). SARS-CoV-2 genomic and replicating RNA, and TMPRSS2 RNA was also detected in the endothelium adjacent to emboli within vessels (Figure 4C).

In the patients with pulmonary metastases from breast cancer and papillary thyroid cancer, some of the cancer cells were positive for SARS-CoV-2 genomic and replicating RNA, ACE2, and TMPRSS2 RNA (Supplementary Figure 2B-2C). In one lung sample, giant cells were positive for SARS-CoV-2 genomic and replicating RNA, and few were positive for ACE2 RNA (Supplementary Figure 2D). FISH also indicated some neutrophils to be positive for SARS-CoV-2 genomic and replicating RNA, ACE2, and TMPRSS2 RNA (Supplementary Figure 2E). Furthermore, hyaline membranes were positive for SARS-CoV-2 genomic and replicating RNA (Figure 4D), likely originating from detached pneumocytes. Finally, we showed that SARS-CoV-2 genomic and replicating RNA, and TMPRSS2 RNA were found in tracheal epithelial cells (Figure 4F).

### SARS-CoV-2, ACE2, and TMPRSS2 in the heart, lymphoid organs, and urogenital tract

Using PCR, we detected SARS-CoV-2 RNA in six of the eight patients in the heart (Figure 2). We found both SARS-CoV-2 genomic and replicating RNA to be present in cardiomyocytes showing that the virus can infect and replicate in cardiomyocytes (Figure 5A). ACE2 was mainly expressed in the stromal cells, and capillary endothelial cells, whereas TMPRSS2 expression was mainly found in perivascular cells and some cardiomyocytes (Figure 5A).

In tonsils, we detected SARS-CoV-2 genomic and replicating RNA in the epithelium, which also expressed ACE2 and TMPRSS2 (Figure 5B; lower panel). Some lymphocytes also showed replicating SARS-CoV-2 RNA (Figure 5B; upper panel). We also observed that ACE2 and TMPRSS2 RNA were expressed by some submucosal cells. In some endothelial cells, we detected only TMPRSS2 RNA but not ACE2 RNA (Figure 5B; upper panel).

In both perihilar and mesenteric lymph nodes, we observed most mesenchymal stromal cells and immune cells to be positive for SARS-CoV-2 genomic and replicating RNA (Figure 5C; upper panel). Some endothelial cells were also infected by the virus (Figure 5C; SARS-CoV-2 genomic RNA, upper panel). Additionally, both ACE2 and TMPRSS2 RNA were expressed strongly in the immune cells within the lymph nodes.

In kidneys, we detected SARS-CoV-2 in different epithelial cells, i.e., glomerular podocytes and parietal epithelial cells (Figure 6A; SARS-CoV-2 genomic RNA, upper panel) and tubular epithelial cells (Figure 6A; lower panel). Similar to lungs, renal endothelium was also infected by the virus, both in glomerular and peritubular capillaries (data not shown).

We detected SARS-CoV-2 also in the prostate, localizing in most prostatic glands, particularly in the glandular epithelial cells (Figure 6B; SARS-CoV-2 genomic and replicating RNA, upper panel). Some mesenchymal cells (Figure 6B, SARS-CoV-2 genomic RNA, bottom panel) and endothelial cells were also infected. ACE2 RNA expression was also found in most glandular epithelial cells (Figure 6B; upper panel) and stromal cells. TMPRSS2 RNA was mainly expressed in the glandular epithelium and stromal cells (Figure 5B) and to a much lesser extent in the endothelial cells.

By RT-PCR, we detected a high viral load of SARS-CoV-2 in the testicles. FISH staining indicated SARS-CoV-2 genomic and replicating RNA to be localized mainly in the germinal epithelium (Figure 6C). Some endothelial cells were also infected by the virus. We observed ACE2 RNA was mainly expressed in the Leydig cells and some endothelial cells, whereas TMPRSS2 was mainly found in the stromal cells. SARS-CoV-2 antisense RNA could also be found in stromal cells of the ovary in one COVID-19 patient (Figure 6D), where ACE2 RNA was also mainly expressed, and some stromal cells expressed TMPRSS2 RNA.

### SARS-CoV-2, ACE2, and TMPRSS2 in the endocrine and gastrointestinal organs, skeletal muscle, and skin

Within the endocrine system, there were six infected thyroids within our autopsy cohort. We detected SARS-CoV-2 genomic and replicating RNA in most follicular epithelial cells (Figure 7A; upper panel), some stromal and endothelial cells (Figure 7A; bottom panel). ACE2 RNA was mainly found in the follicular epithelium (Figure 7A; upper panel) and endothelial cells, whereas TMPRSS2 RNA was expressed in most follicular epithelial cells (Figure 7A; upper panel), some stromal and endothelial cells.

In the two adrenal glands, we observed SARS-CoV-2 genomic and replicating SARS-CoV-2 RNA in most glandular epithelial cells (Figure 7B) and to a lesser extent in stromal and endothelial cells. ACE2 RNA expression was mainly found in most endothelial cells. TMPRSS2 RNA was mainly expressed in glandular epithelial cells (Figure 7B) and some endothelial cells.

In the salivary gland, SARS-CoV-2 genomic and replicating RNA were detected in most acini and salivary ducts together with strong detection of ACE2 and TMPRSS2 RNA (Figure 7C). SARS-CoV-2 genomic RNA was also found in some stromal cells and endothelial cells. ACE2 RNA expression was found in some stromal cells and most endothelial cells but TMPRSS2 RNA was not found in these two cell types.

Within the small bowel, SARS-CoV-2 genomic and replicating SARS-CoV-2 RNA were mainly detected in the enterocytes, accompanied by strong expression of ACE2 and TMPRSS2 RNA (Figure 7C). Additionally, ACE2 RNA expression was found in mesenchymal stromal cells, muscle cells expressed both ACE2 and TMPRSS2 RNA. In the endothelial cells, we detected ACE2 but not TMPRSS2 RNA.

In the pancreas, we detected genomic and replicating SARS-CoV-2 RNA in most acinar cells, which colocalized with strong expression of ACE2 and TMPRSS2 RNA (Figure 7D). SARS-CoV-2 genomic RNA was also detectable in some stromal and endothelial cells. ACE2 RNA was present in most stromal cells and some endothelial cells, while TMPRSS2 RNA was only present in some stromal cells.

We detected genomic and replicating SARS-CoV-2 RNA in the skeletal muscle, mainly localized in the myocytes (Figure 7E) and some endothelial cells. Moreover, the muscle cells also expressed ACE2 RNA and TMPRSS2 RNA. In the endothelial cells, we observed ACE2 RNA expression but not TMPRSS2 RNA.

Finally, we detected both SARS-CoV-2 genomic and replicating RNA in the dermal epithelial cells (Figure 7F). The epithelial cells also expressed ACE2 and TMPRSS2, while mRNA for both proteins was only sparsely detected in endothelial cells.

### IHC of SARS-CoV-2 spike glycoprotein on FFPE autopsy lung samples

Protein detection of SARS-CoV-2 spike glycoprotein is widely used in studies in FFPE tissues. We stained for the SARS-CoV-2 spike glycoprotein in the COVID-19 autopsy lung and selected the most commonly used antibodies based on previous studies ^19,22^, targeting the SARS spike glycoprotein (Abcam, ab272420) and SARS-CoV SΔ10 within S2 domain protein (Genetex, GTX632604). We have tested these two antibodies on FFPE autopsy lung tissues with four different antigen retrieval methods as outlined in the methods section. All approaches were unsuccessful, i.e., no positive signals could be observed, except the ab272420 using Tris-EDTA buffer (pH 9) antigen retrieval. We validated this protocol on autopsy lung tissues from our COVID-19 cohort and non-COVID-19 cases as negative controls, including 1) four patients infected with influenza, 2) six patients who developed ARDS (selected non-superinfected areas), and 3) six patients who had no pulmonary pathology (Table 2). These autopsy tissues were collected before the COVID-19 outbreak (from 2009 to July 2019). All lung tissues showed variable but very distinct false-positive unspecific stain, except for a single lung from the non-infected group (Table 2, Figure 8, Supplementary Figure 3). Taken together, IHC based detection of SARS-CoV-2 using the two mentioned antibodies and protocols were not possible to establish or not specific to detect the virus in FFPE tissues.

**Figure 8.**
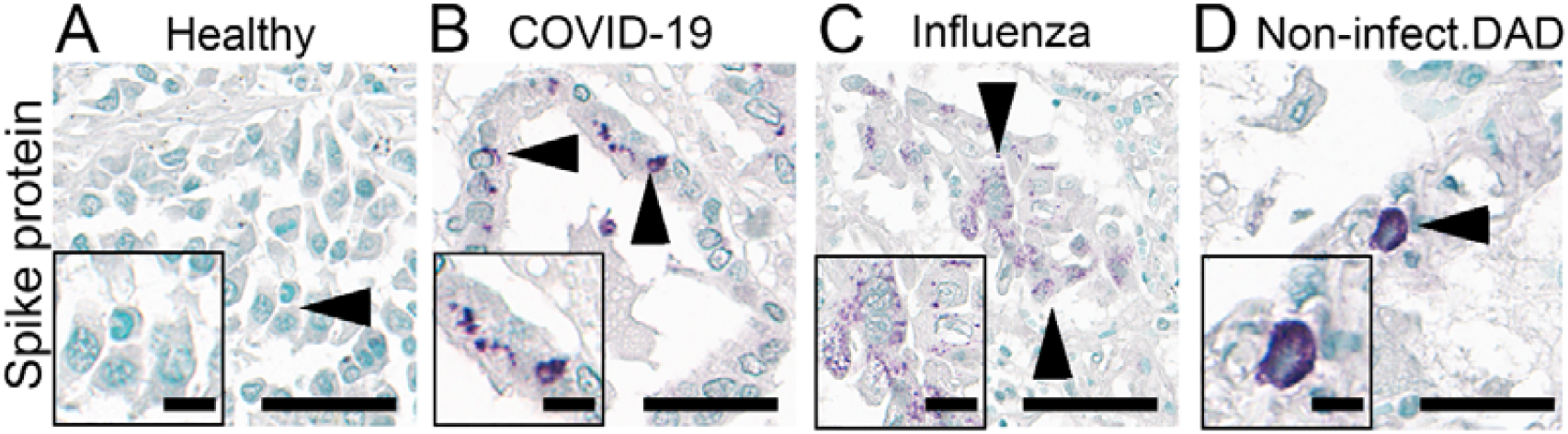
Immunohistochemical staining of SARS spike glycoprotein. IHC staining of SARS spike glycoprotein (#Ab272420) in the lung autopsies collected from patients without any respiratory disease (Healthy; insert: respiratory epithelial cells) and patients infected with COVID-19 (insert: respiratory epithelial cells), influenza (insert: respiratory epithelial cells) and non-infectious diffuse alveolar damage (DAD; insert: cell with features of an alveolar macrophage) with apparently (false-) positive staining (arrowhead). Scale bars represent 40 µm and 10 µm (insert).

### Visualization of SARS-CoV-2 by electron microscopy

Ultrastructural tissue preservation in autopsy material was rather poor and hampered virus particle detection by electron microscopy. Autolytic processes before fixation led to cell degradation and strong autolytic vesicle formation, resulting in many virus-like particles, but not a single reliable SARS-CoV-2 detection, despite a thorough analysis of all cases (data not shown).

### Application of chromogenic *in situ* hybridization on external FFPE COVID-19 tissue samples

FISH requires fluorescence microscopes, which might not be available in all pathologies performing autopsies, and the signal also faints with time. Therefore, we also established the chromogenic *in situ* hybridization (CISH) for SARS-CoV-2 detection (Supplementary method, Supplementary table 3). We first validated the CISH detection with the same positive and negative control probes used in FISH (Supplementary Figure 4A-D). To validate the compatibility of our ISH detection of SARS-CoV-2 in FFPE autopsies processed externally, we performed CISH staining on an independent autopsy cohort consisting of nine COVID-19 patients received from the Institute of Pathology, Hannover Medical School, Germany (Supplementary Figure 5). We could detect SARS-CoV-2 genomic and replicating RNA in the cardiomyocytes, intra-glomerular and alveolar cells, hepatocytes, pancreatic and enteric epithelial cells, capillary endothelial cells in lymph nodes, and spleen (Supplementary figure 5A-I). In the vicinity of the SARS-CoV-2 positive signals, we also detected ACE2 and TMPRSS2 RNA in intra-glomerular cells, hepatocytes, alveolar cells, pancreatic and enteric epithelial cells.

## Discussion

We here comprehensively analyzed SARS-CoV-2 in tissues from clinical autopsies of COVID-19 cases. We compared various methods, suggesting the most effective and useful approaches for SARS-CoV-2 detection in autopsy tissues, focusing on routine processing in pathology and thereby possibly the most wide spread adoption of the methods. Using these methods, we have comprehensively analyzed a multitude of tissues and cells for SARS-CoV-2 presence to perform a detailed correlation of virus presence and potential pathology findings. We describe virus spread in several tissues (i.e., thyroid, adrenal gland, prostate, large bowel and ovary) previously not described and pin-point the localization to specific cells within each tissue.

Comparing various approaches routinely used in pathology institutes, the RNA detection methods (either by RT-PCR, FISH or CISH) were proved to be the most effective, specific, and suitable to use. In contrast, both immunohistochemistry and electron microscopy turned out not to be suitable, confirming previous reports ^11,23,24^. When using RNA-based methods, a 2-step approach could be applied. First, screening the cases and tissues using the RT-PCR, followed by a more complex and time-consuming FISH approach only in positive tissues. We focused our approach on FFPE tissue since this is internationally the most widely adopted tissue processing in institutes of pathology and autopsy centers. Although formalin fixation and paraffin embedding might lead to additional RNA degradation, FFPE material has the important advantage of not being infectious and conservable, which makes further processing much easier and more broadly applicable compared to, e.g., fresh infectious tissue. Further, reliable detection in FFPE materials allows for fast retrospective analysis, as FFPE blocks are regularly archived and widely available in all institutes of pathology.

Compared to RT-PCR, the ISH detection method in FFPE autopsies could avoid the interference of SARS-CoV-2 RNA in the blood that remained within the tissues ^9^. To precisely localize the signal, consecutive sections can be used, allowing a very detailed association of morphology with positive signals, which is not possible in FISH or CISH sections alone. Apart from the precise localization, the FISH approach also allows colocalizing analysis of the viral RNA and its targets ACE2 and TMPRSS2 on the cellular level. Compared to FISH, the CISH method has some advantages, particularly not requiring fluorescence microscopy equipment, which might not be readily available in all institutes, longer stability of the signals, and the possibility of a faster digitalization of the slides using whole-slide scanners. On the other hand, the FISH signals are easier to detect and provide a much better signal-to-noise ratio by adjusting the fluorescence excitation. Validation of the ISH on an independent external cohort in our study confirmed its broad applicability.

The macroscopic and microscopic findings of the autopsy lungs are fully in line with previous reports on pulmonary and upper airway findings in COVID-19 ^11,20,25,26^ and SARS-CoV-1 autopsies ^27,28^. The strong positivity for viral RNA, including the antisense, as well as the co-localization with ACE2 and TMPRSS2 associated the pathological findings with the viral presence and are also in line with previous reports. We here provided comprehensive morpho-molecular analyses and examples of the localization of the virus in various cells and compartments of the lung (i.e., squamous metaplasia and giant cell), which were previously not reported.

Currently, COVID-19 is considered a multisystemic disease. Our data, showing the viral presence, including replicating virus, in the majority of analyzed tissues, might confirm this hypothesis. However, in our macroscopic and microscopic evaluation of all extra-respiratory organs, we were not able to identify specific pathological alterations that would be attributable to viral infection. We cannot exclude subtle or molecular changes that were not translating to the morphological changes presented in the infected non-respiratory tissues. However, our detailed analyses might suggest that the SARS-CoV-2 spreading beyond the respiratory tract does not induce any major pathology and might be rather negligible in comparison to the pulmonary involvement, at least in fatal cases. Similar conclusions were also suggested in some previous reports ^10,20,29^.

One hypothesis for the multisystemic nature of COVID-19 is the systemic spread and effects on the (micro)vasculature ^30^. We show SARS-CoV-2 genomic and replicating RNA and particularly TMPRSS2 RNA in the endothelial cells of vessels across all organs, supporting this hypothesis. This, is supported by data showing SARS-CoV-2 RNA in the blood ^31^. Besides, we also found the viral RNA in vessels across all organ systems and pulmonary endothelial cells adjacent to thrombi, which might be a hint to the microvascular damage induced by direct endothelial infection^10^. However, we have only found vasculature with thrombi in the lungs, but not in any other of the analyzed organs or tissues. This might be influenced by our relatively small cohort, mainly consisting of intensive care unit patients who had been receiving anticoagulative treatment. We have also been very cautious regarding the interpretation of vascular occlusion as intravital thrombotic events in order not to false-positively interpret post-mortem clots as intravital thrombi.

We showed the predominance of viral RNA in ACE2 and/or TMPRSS2-expressing epithelial cells compared to mesenchymal stromal cells and endothelial cells in various organs. This included e.g., trachea, lung, tonsil, salivary glands, kidney, and small bowel. The presence of SARS-CoV-2 RNA in salivary glands supports the effectiveness of saliva-based SARS-CoV-2 testing that has been emerged worldwide ^32^. We also showed the presence of viral RNA in non-epithelial tissues, such as lymph nodes, heart, and skeletal muscle ^20,33-35^. We visualized for the first time SARS-CoV-2 viral RNA in exo/endocrine organs, i.e., in the thyroid, adrenal gland, prostate, testicle, and ovary. Our findings in testicle are in line with previous reports of detection of SARS-CoV-2 RNA in seminal fluid and testicular tissue with RT-PCR ^36,37^. Regarding adrenal glands and prostate in COVID-19, microinfarctions, hemorrhage or thrombosis have been described in COVID-19 autopsies ^38,39^. Concerning the thyroid, previous descriptions of follicular epithelial morphologic alterations exist for SARS infection ^40^, but not COVID-19. We also detected SARS-CoV-2 RNA in cancer cells in two cases of pulmonary metastasis. This finding is in line with a previous report of a positive RT-PCR result in oral cancer tissue of a COVID-19 patient ^41^. Current knowledge on the potential impact of SARS-CoV-2 infection in cancer cells remains incomplete, beyond the observation of their higher morbidity and mortality of cancer patients after being infected with the SARS-CoV-2 virus ^42^.

We could also confirm the presence of ACE2 receptor on alveolar epithelial cells ^43^ and pancreatic ductal cells ^44^ as well as TMPRSS2 on salivary acinar cells ^45^ and prostate epithelial cells ^46^ as previously suggested by single-cell RNA sequencing studies.

Our study has several limitations. We have not found viral presence in a few of the analyzed tissues, including bone marrow, esophagus, large bowel, and spleen. This might be due to the low number of analyzed patients. We aimed to perform a detailed analysis and cellular localization, which in the absence of reliable protein or ultrastructural detection, required to perform FISH analysis. This method is time-consuming, particularly when using co-localization with consecutive sections, making it hard to apply in a large number of cases. Therefore, for more high-throughput approaches, we suggest pre-screening of the cases and tissues using RT-PCR.

Our study is descriptive, provides a single time-point analysis of only fatal cases. However, these are intrinsic limitations of autopsy studies. On the other hand, there are no other approaches that would allow analyzing comprehensively all human tissues in a comparable way. The variable degrees of autolysis in some tissue samples might limit the efficacy of viral detection. This was particularly true for electron microscopy, which was not applicable in our hands. Methodologically, in some cell types with scarce cytoplasm and overlap with neighbouring cells, it was not possible to allocate the ISH signal localization to a particular cell. We found a small population of SARS-CoV-2 positive cells that were negative for ACE2 or TMPRSS2 RNA. This is most likely due to a sampling bias, given that the FISH or CISH slides are extremely thin and only reflect part of the volume of the cells. An alternative explanation might be that SARS-CoV-2 was suggested to enter host cells independently of ACE2 and TMPRSS2 ^47^. Regarding the wide spread of SARS-CoV-2 variants lately, it is unclear whether our RNA-targeted approaches would detect or being interfered by the mutations, despite the fact that a pool of probes for detecting different genomic sites of the SARS-CoV-2 is used in our FISH method.

In conclusion, our present study suggests that RT-PCR and FISH performed on FFPE tissues are reliable methods allowing tissue and cellular localization in autopsies. We provide a “cellular and whole body tissue atlas” of organ tropism profile of SARS-CoV-2 in severe COVID-19 patients, suggesting a multisystemic spread in virtually all cell types of infected organs.

## Data Availability

The datasets generated and analyzed within the current study are not publicly available due to protecting the study participant privacy but are available from the corresponding author on reasonable request.

## Author contributions

DWLW and MCT performed FISH, CISH stains, DWLW, BMK and SD performed FISH analysis. DWLW and SD performed immunohistochemistry analysis. DWLW, SD, and BMK created figures. DWLW, SD, BMK and SVS wrote the manuscript. EB performed electron microscopy. RDB, SVS, and SW performed autopsies and histological analyses. RKC, CC, TB, and PB supervised autopsy assessment. SV and ED performed PCR analyses. PB had the conceptual idea. All authors read and approved the final version of the article.

## Acknowledgments

The excellent technical help of Inge Losen and Simon Otten is gratefully acknowledged. We wish to thank all relatives and legal representatives for giving their consent for the autopsy procedure.

## Supplementary Material

### Supplementary Method

#### External study cohort

Supplementary Table 1. List of autopsies from different decedents organized in the TMA.

Supplementary Table 2. Major pathological findings in the autopsy cohort.

Supplementary Table 3. Patient characteristics of the external COVID-19 study cohort.

Supplementary Figure 1. Validation of FISH detection on human autopsies.

Supplementary Figure 2. Virus detection in respiratory system by FISH.

Supplementary Figure 3. Virus detection in respiratory system by IHC staining.

Supplementary Figure 4. Validation of CISH detection on external human autopsies.

Supplementary Figure 5. Detection of SARS-CoV-2 sense, antisense, ACE2 and TMPRSS2 on an external sample by CISH.

## References

1. Cabrera Martimbianco AL, Pacheco RL, Bagattini AM, Riera R. Frequency, signs and symptoms, and criteria adopted for long COVID: a systematic review. Int J Clin Pract 2021: e14357.

2. Wei JF, Huang FY, Xiong TY, et al. Acute myocardial injury is common in patients with COVID-19 and impairs their prognosis. Heart 2020; 106(15): 1154–9.

3. Maiese A, Manetti AC, La Russa R, et al. Autopsy findings in COVID-19-related deaths: a literature review. Forensic Sci Med Pathol 2020.

4. Rensen E, Pietropaoli S, Weber C, et al. Sensitive visualization of SARS-CoV-2 RNA with CoronaFISH. bioRxiv 2021: 2021.02.04.429604.

5. Gagiannis D, Umathum V, Bloch W, et al. Antemortem vs. postmortem histopathological and ultrastructural findings in paired transbronchial biopsies and lung autopsy samples from three patients with confirmed SARS-CoV-2 infection. medRxiv 2021: 2020.12.30.20248929.

6. Shieh WJ, Hsiao CH, Paddock CD, et al. Immunohistochemical, in situ hybridization, and ultrastructural localization of SARS-associated coronavirus in lung of a fatal case of severe acute respiratory syndrome in Taiwan. Hum Pathol 2005; 36(3): 303–9.

7. Ackermann M, Mentzer SJ, Jonigk D. Visualization of SARS-CoV-2 in the Lung. Reply. N Engl J Med 2020; 383(27): 2689–90.

8. Bharat A, Querrey M, Markov NS, et al. Lung transplantation for patients with severe COVID-19. Sci Transl Med 2020; 12(574).

9. Bhatnagar J, Gary J, Reagan-Steiner S, et al. Evidence of Severe Acute Respiratory Syndrome Coronavirus 2 Replication and Tropism in the Lungs, Airways, and Vascular Endothelium of Patients With Fatal Coronavirus Disease 2019: An Autopsy Case Series. J Infect Dis 2021; 223(5): 752–64.

10. Bussani R, Schneider E, Zentilin L, et al. Persistence of viral RNA, pneumocyte syncytia and thrombosis are hallmarks of advanced COVID-19 pathology. EBioMedicine 2020; 61: 103104.

11. Massoth LR, Desai N, Szabolcs A, et al. Comparison of RNA In Situ Hybridization and Immunohistochemistry Techniques for the Detection and Localization of SARS-CoV-2 in Human Tissues. Am J Surg Pathol 2021; 45(1): 14–24.

12. Schurink B, Roos E, Radonic T, et al. Viral presence and immunopathology in patients with lethal COVID-19: a prospective autopsy cohort study. Lancet Microbe 2020; 1(7): e290–e9.

13. Bouquegneau A, Erpicum P, Grosch S, et al. COVID-19-associated nephropathy includes tubular necrosis and capillary congestion, with evidence of SARS-CoV-2 in the nephron. Kidney360 2021: 10.34067/KID.0006992020.

14. Pfister F, Vonbrunn E, Ries T, et al. Complement Activation in Kidneys of Patients With COVID-19. Front Immunol 2020; 11: 594849.

15. Best Rocha A, Stroberg E, Barton LM, et al. Detection of SARS-CoV-2 in formalin-fixed paraffin-embedded tissue sections using commercially available reagents. Laboratory investigation; a journal of technical methods and pathology 2020; 100(11): 1485–9.

16. Hoffmann M, Kleine-Weber H, Schroeder S, et al. SARS-CoV-2 Cell Entry Depends on ACE2 and TMPRSS2 and Is Blocked by a Clinically Proven Protease Inhibitor. Cell 2020; 181(2): 271–80 e8.

17. Wang F, Flanagan J, Su N, et al. RNAscope: a novel in situ RNA analysis platform for formalin-fixed, paraffin-embedded tissues. J Mol Diagn 2012; 14(1): 22–9.

18. Klinkhammer BM, Djudjaj S, Kunter U, et al. Cellular and Molecular Mechanisms of Kidney Injury in 2,8-Dihydroxyadenine Nephropathy. Journal of the American Society of Nephrology : JASN 2020; 31(4): 799–816.

19. von Stillfried S, Villwock S, Bulow RD, et al. SARS-CoV-2 RNA screening in routine pathology specimens. Microb Biotechnol 2021.

20. Remmelink M, De Mendonca R, D’Haene N, et al. Unspecific post-mortem findings despite multiorgan viral spread in COVID-19 patients. Crit Care 2020; 24(1): 495.

21. Sato S, Uejima Y, Arakawa Y, et al. Clinical features of macrophage activation syndrome as the onset manifestation of juvenile systemic lupus erythematosus. Rheumatol Adv Pract 2019; 3(1): rkz013.

22. Bradley BT, Maioli H, Johnston R, et al. Histopathology and ultrastructural findings of fatal COVID-19 infections in Washington State: a case series. Lancet 2020; 396(10247): 320–32.

23. Hopfer H, Herzig MC, Gosert R, et al. Hunting coronavirus by transmission electron microscopy - a guide to SARS-CoV-2-associated ultrastructural pathology in COVID-19 tissues. Histopathology 2021; 78(3): 358–70.

24. Smith KD, Akilesh S, Alpers CE, Nicosia RF. Am I a coronavirus? Kidney Int 2020; 98(2): 506–7.

25. Polak SB, Van Gool IC, Cohen D, von der Thusen JH, van Paassen J. A systematic review of pathological findings in COVID-19: a pathophysiological timeline and possible mechanisms of disease progression. Mod Pathol 2020; 33(11): 2128–38.

26. Bosmuller H, Traxler S, Bitzer M, et al. The evolution of pulmonary pathology in fatal COVID-19 disease: an autopsy study with clinical correlation. Virchows Arch 2020; 477(3): 349–57.

27. Hwang DM, Chamberlain DW, Poutanen SM, Low DE, Asa SL, Butany J. Pulmonary pathology of severe acute respiratory syndrome in Toronto. Mod Pathol 2005; 18(1): 1–10.

28. Nicholls JM, Poon LL, Lee KC, et al. Lung pathology of fatal severe acute respiratory syndrome. Lancet 2003; 361(9371): 1773–8.

29. Matschke J, Lutgehetmann M, Hagel C, et al. Neuropathology of patients with COVID-19 in Germany: a post-mortem case series. Lancet Neurol 2020; 19(11): 919–29.

30. Ackermann M, Verleden SE, Kuehnel M, et al. Pulmonary Vascular Endothelialitis, Thrombosis, and Angiogenesis in Covid-19. N Engl J Med 2020; 383(2): 120–8.

31. Buetti N, Patrier J, Le Hingrat Q, et al. Risk factors for SARS-CoV-2 detection in blood of critically ill patients. Clin Infect Dis 2020.

32. Tan SH, Allicock O, Armstrong-Hough M, Wyllie AL. Saliva as a gold-standard sample for SARS-CoV-2 detection. The Lancet Respiratory Medicine.

33. Huang N, Perez P, Kato T, et al. SARS-CoV-2 infection of the oral cavity and saliva. Nat Med 2021.

34. Puelles VG, Lutgehetmann M, Lindenmeyer MT, et al. Multiorgan and Renal Tropism of SARS-CoV-2. N Engl J Med 2020; 383(6): 590–2.

35. Sekulic M, Harper H, Nezami BG, et al. Molecular Detection of SARS-CoV-2 Infection in FFPE Samples and Histopathologic Findings in Fatal SARS-CoV-2 Cases. Am J Clin Pathol 2020; 154(2): 190–200.

36. Li D, Jin M, Bao P, Zhao W, Zhang S. Clinical Characteristics and Results of Semen Tests Among Men With Coronavirus Disease 2019. JAMA Netw Open 2020; 3(5): e208292.

37. Yang M, Chen S, Huang B, et al. Pathological Findings in the Testes of COVID-19 Patients: Clinical Implications. Eur Urol Focus 2020; 6(5): 1124–9.

38. Hanley B, Naresh KN, Roufosse C, et al. Histopathological findings and viral tropism in UK patients with severe fatal COVID-19: a post-mortem study. Lancet Microbe 2020; 1(6): e245–e53.

39. Wichmann D, Sperhake JP, Lutgehetmann M, et al. Autopsy Findings and Venous Thromboembolism in Patients With COVID-19: A Prospective Cohort Study. Ann Intern Med 2020; 173(4): 268–77.

40. Wei L, Sun S, Xu CH, et al. Pathology of the thyroid in severe acute respiratory syndrome. Hum Pathol 2007; 38(1): 95–102.

41. Guerini-Rocco E, Taormina SV, Vacirca D, et al. SARS-CoV-2 detection in formalin-fixed paraffin-embedded tissue specimens from surgical resection of tongue squamous cell carcinoma. J Clin Pathol 2020; 73(11): 754–7.

42. van Dam PA, Huizing M, Mestach G, et al. SARS-CoV-2 and cancer: Are they really partners in crime? Cancer Treat Rev 2020; 89: 102068.

43. Sungnak W, Huang N, Becavin C, et al. SARS-CoV-2 entry factors are highly expressed in nasal epithelial cells together with innate immune genes. Nat Med 2020; 26(5): 681–7.

44. Kusmartseva I, Wu W, Syed F, et al. Expression of SARS-CoV-2 Entry Factors in the Pancreas of Normal Organ Donors and Individuals with COVID-19. Cell Metab 2020; 32(6): 1041–51 e6.

45. Song J, Li Y, Huang X, et al. Systematic analysis of ACE2 and TMPRSS2 expression in salivary glands reveals underlying transmission mechanism caused by SARS-CoV-2. Journal of medical virology 2020; 92(11): 2556–66.

46. Song H, Seddighzadeh B, Cooperberg MR, Huang FW. Expression of ACE2, the SARS-CoV-2 receptor, and TMPRSS2 in prostate epithelial cells. bioRxiv 2020.

47. Cantuti-Castelvetri L, Ojha R, Pedro LD, et al. Neuropilin-1 facilitates SARS-CoV-2 cell entry and infectivity. Science 2020; 370(6518): 856–60.

