## Supplementary Material for "Multisystemic cellular tropism of SARS-CoV-2 in autopsies of COVID-19 patients"

Running Title: Cell atlas of COVID infection

Dickson W.L. Wong^1,*^, Barbara M. Klinkhammer^1,*^, Sonja Djudjaj^1,*^, Sophia Villwock^1^, M. Cherelle Timm^1^, Eva M. Buhl^1,2^, Sophie Wucherpfennig^1^, Claudio Cacchi^1^, Till Braunschweig^1^, Ruth Knüchel-Clarke^1^, Danny Jonigk^4,5^, Christopher Werlein^4^, Roman D. Bülow^1^, Edgar Dahl^1^, Saskia von Stillfried^1,*^, Peter Boor ^1,2,3,*^

^1^ Institute of Pathology, RWTH Aachen University Hospital, Aachen, Germany

^2^ Electron Microscopy Facility, RWTH Aachen University Hospital, Aachen, Germany

^3^ Department of Nephrology and Immunology, RWTH Aachen University Hospital, Aachen, Germany

^4^ Institue of Pathology, Hannover Medical School, Hannover, Germany

^5^ Member of the German Center for Lung Research (DZL), Biomedical Research in Endstage and Obstructive Lung Disease Hannover (BREATH)

* Authors contributed equally

**Supplementary information**

Supplementary Method

External study cohort

Supplementary Table 1. List of autopsies from different decedents organized in the TMA.

Supplementary Table 2. Major pathological findings in the autopsy cohort.

Supplementary Table 3. Patient characteristics of the external COVID-19 study cohort.

Supplementary Figure 1. Validation of FISH detection on human autopsies.

Supplementary Figure 2. Virus detection in respiratory system by FISH.

Supplementary Figure 3. Virus detection in respiratory system by IHC staining.

Supplementary Figure 4. Validation of CISH detection on external human autopsies.

Supplementary Figure 5. Detection of SARS-CoV-2 sense, antisense, ACE2 and TMPRSS2 on an external sample by CISH.

**Supplementary Method**

**SARS-CoV-2 RNA detection with Chromogenic *in situ* hybridization (CISH)**

We performed CISH staining on the 1-μm-thick paraffin tissue microarray sections (n = 9 patients) received from an external COVID-19 autopsy center (provided by Prof. D. Jonigk, Institute of Pathology, Hannover Medical School) with the RNAscope® 2.5 HD Duplex Reagent Kit (Advanced Cell Diagnostics, Inc., Hayward, California). The procedure is analogous to FISH until the amplifier steps. Instead of fluorophore, horseradish peroxidase (HRP) and alkaline phosphatase (AP) were conjugated to the C1 (cyan) and C2 (red) probe, respectively. We applied the same probes used in FISH staining to the tissues. Lastly, tissue slides were counterstained with Gills Haematoxylin (Sigma-Aldrich, St. Louis, MO) and mounted with EcoMount solution (Biocare Medical; Concord, California). Stained slides were analyzed with Aperio VERSA Brightfield, Fluorescence & FISH Digital Pathology Scanner (Leica Microsystems Inc, Buffalo Grove, IL, USA).

**External study cohort**

The main characteristics of the external study cohort (9 male/ 4 female; median age 76 [54–96 years) are given in Table XX. The period between the hospital admission and death ranged from 3 to 22 days (median 9 days, Figure 1). 12 patients had a previous diagnosis of hypertension (n=11) and/ or diabetes mellitus type 2 (n=5). Four patients had a history of tobacco abuse, four patients were non-smokers, while no information on current or previous tobacco abuse was available for the remaining five patients. Six patients were treated with mechanical ventilation. One patient refused mechanical ventilation. Five patients received no ventilation therapy due to a palliative situation.

**Supplementary Table 1. List of autopsies from different decedents organized in the TMA blocks.** Autopsies were processed with RT-PCR (P) and FISH staining (F). Tissues that were not available are labeled as (N/A).

| **Patient/ Sex** | **1/ M** | **2/ F** | **3/ M** | **4/ M** | **5/ M** | **6/ F** | **7/ M** | **8/ F** |
| --- | --- | --- | --- | --- | --- | --- | --- | --- |
| Lung central | P, F | P, F | P, F | P, F | P, F | P, F | P, F | P, F |
| Lung periphery | P, F | P, F | P, F | P, F | P, F | P, F | P, F | P, F |
| Distal trachea | P, F | P, F | P, F | P, F | P, F | P, F | P, F | P, F |
| Heart | P, F | P, F | P, F | P, F | P, F | P, F | P, F | P, F |
| Kidney | P, F | P, F | P, F | P, F | P, F | P, F | F | P, F |
| Liver | P, F | P, F | P, F | P, F | P, F | P, F | P, F | P, F |
| Adrenal gland | P, F | P, F | P, F | P, F | P, F | P, F | P, F | P, F |
| Spleen | P, F | P, F | P, F | P, F | P, F | P, F | P, F | P, F |
| Small bowel | F | F | N/A | F | F | F | F | F |
| Large bowel | F | F | F | P, F | F | F | F | F |
| Lymph node (perihilar) | P, F | P, F | P, F | P, F | P, F | P, F | P, F | F |
| Salivary gland | P, F | P, F | P, F | P, F | P, F | P, F | P, F | P, F |
| Tonsil | P, F | P, F | P, F | P, F | P, F | P, F | P, F | P |
| Bone Marrow | P | P | N/A | P | P, F | N/A | N/A | N/A |
| Ovary | N/A | P, F | N/A | N/A | N/A | N/A | N/A | F |
| Testicles | P, F | N/A | P, F | P, F | F | N/A | P, F | P, F |
| Uterus | N/A | F | N/A | N/A | N/A | P, F | N/A | P, F |
| Prostate | P, F | N/A | P, F | P, F | F | N/A | F | N/A |
| Urinary bladder | N/A | N/A | N/A | N/A | F | N/A | N/A | N/A |
| Oropharynx | P, F | P, F | F | P, F | P, F | P, F | P, F | P, F |
| Esophagus | P, F | P, F | P | P, F | P, F | P, F | P, F | P, F |
| Stomach | P, F | P, F | P, F | P, F | P, F | P, F | P, F | P, F |
| Pancreas | P, F | P, F | P, F | P, F | P, F | P, F | P, F | P, F |
| Thyroid | P, F | P, F | P, F | P, F | P, F | P, F | P, F | P, F |
| Muscle | P, F | F | P, F | P, F | F | P, F | P, F | P |
| Skin | P | F | P, F | P, F | N/A | P, F | P, F | P, F |
| Lymph node (mesenteric) | P, F | P, F | P, F | P, F | P, F | P, F | N/A | P |

**Supplementary Table 2. Major pathological findings in the autopsy cohort.**

|  | Pulmonary | | | | | | Extra-pulmonary | | |
| --- | --- | --- | --- | --- | --- | --- | --- | --- | --- |
|  |  |  |  |  |  |  | Cardiovascular | Urogenital | Gastrointestinal |
|  |  | | | | | | Heart | Kidney | Liver |
| Patient/ sex | DAD | Inflammation | Giant cells | Squamous metaplasia | Hyaline membranes | Additional observation |  |  |  |
| 1/ M | Proliferative/ organizing phase | Interstitial infiltrate | + | + | + | Intraalveolar edema, focal organized pneumonia | Normal heart weight, fibrosis in endocardium, patchy myocardium infiltrate | Type 1 diabetes and nodular glomerulosclerosis, no significant chronic inflammation | Steatosis, intracytoplasmic cholestasis |
| 2/ F | Proliferative/ organizing phase | Interstitial lymphocytic inflammation | - | ++ | + | Hyaline microthrombi, | Slight cardiomegaly, patchy lymphocytic infiltrate in epicardium, macrophages (signs of previous bleeding), absorptive/ reactive inflammation, endocardial fibrosis, discrete pericarditis, perivascular fibrosis, signs of cellular hypertrophy | Inflammatory cells only in fibrotic areas | Liver cirrhosis, chronic inflammation in capsule and fibrotic areas, steatosis |
| 3/ M | Proliferative phase | Superinfection with acute inflammation | - | - | + (very focal) | Intra-alveolar edema, Intra-alveolar hemorrhage | Cardiomegaly, cellular hypertrophy, very scarce lymphocytes, single cell necrosis, focal fibrosis | Hypertensive damage, inflammation only in fibrotic areas, no peritubular capillaritis | Disseminated inflammation, steatosis |
| 4/ M | Proliferative/ organizing phase | Interstitial inflammation | + | + | - | Hemorrhage, thromboembolus in small pulmonary arteries | Cardiomegaly, perivascular lymphocyte, eosinophilic neutrophil infiltrate, discrete chronic inflammation | Chronic inflammation, single sclerotic glomeruli, acute kidney injury | Cholangitis lenta, chronic inflammation of biliary ducts |
| 5 / M | Proliferative/ organizing | Acute pneumonia with granulocytes, macrophage, acute inflammation | + | + | + | Intraalveolar edema, intra-alveolar hemorrhage | Cardiomegaly, patchy myocardial infiltrate, histiocytic infiltrate, discrete chronic inflammation | Chronic inflammation, single sclerotic glomeruli, acute kidney injury, peritubular capillaries/ vasa recta with lots of leukocytes | Cholangitis lenta, chronic inflammation of biliary ducts |
| 6/ F | Proliferative phase | Interstitial inflammation | ++ | + | + | Thrombus, Lung metastasis, intraalveolar edema, bronchitis | Slight cardiomegaly, patchy myocardial infiltrate | Patchy inflammatory infiltrate, some peritubular capillaries with leukocytes | Lymphocytes in sinusoids, no other pathology |
| 7/ M | Proliferative and fibrotic phase | Intra-alveolar granulocytes | ++ | + | - | Intraalveolar edema, | Normal heart weight, histiocytic infiltrate | Focal chronic inflammation, primarily in fibrotic areas, some in peritubular capillaries | Cholangitis lenta, chronic inflammation of biliary ducts |
| 8/ F | Proliferative and fibrotic phase | Granulocytes, chronic inflammatory infiltrate | - | - | + | Intraalveolar hemorrhage, intraalveolar edema | Slight cardiomegaly, inflammation only in fibrosis | Rejection, cirrhotic kidney | Perivascular and sinusoidal lymphocytes |

**Supplementary Table 3. Patient characteristics of the external COVID-19 study cohort.**

BMI = Body-mass index; d = day; y = year

| Patient | Sex | Age range (y) | BMI | Smoker | Hospital admission to death (d) | Ventilation therapy |
| --- | --- | --- | --- | --- | --- | --- |
| 1 | M | 91-95 | Underweight | No | 14 | No |
| 2 | M | 76-80 | Moderately obese | n.a. | 22 | Yes |
| 3 | M | 71-75 | Severely obese | n.a. | 12 | Yes |
| 4 | F | 66-70 | n.a. | n.a. | 14 | n.a. |
| 5 | F | 76-80 | n.a. | n.a. | 7 | Yes |
| 6 | M | 51-55 | n.a. | n.a. | 21 | Yes |
| 7 | M | 76-80 | Severely obese | Yes | 3 | Yes |
| 8 | M | 66-70 | Overweight | Yes | 9 | Yes |
| 9 | M | 71-75 | Overweight | Yes | 3 | No |
| 10 | F | 81-85 | Overweight | Yes | 4 | No |
| 11 | F | 66-70 | Severely obese | No | 9 | No |
| 12 | M | 86-90 | Overweight | No | 5 | No |
| 13 | M | 96-100 | Normal weight | No | 3 | No |

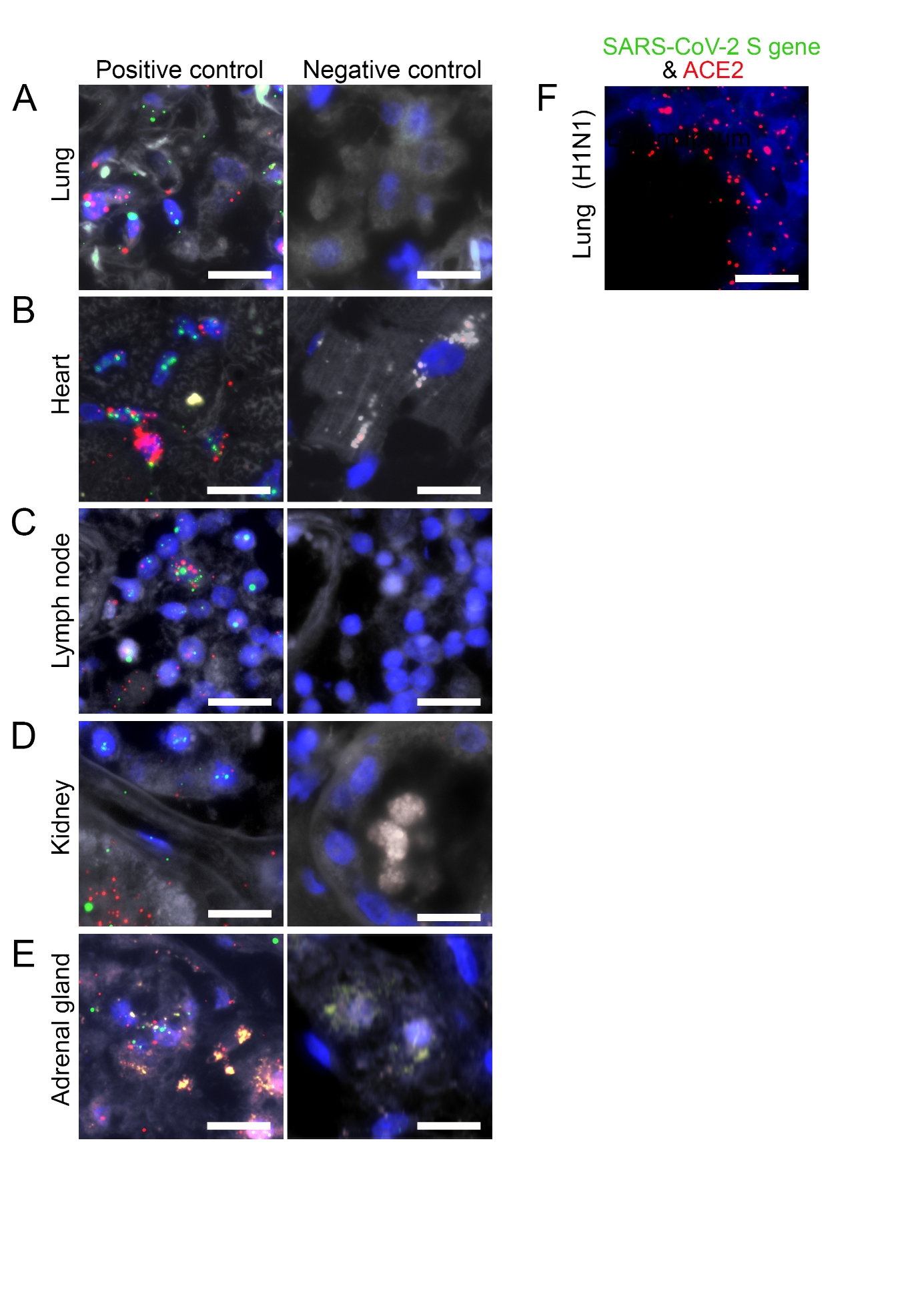

**Supplementary Figure 1. Validation of FISH detection on human autopsies.**

FISH method was validated by incubation of probes targeting *Homo sapiens* *POLR2A* (green) and *PPIB* (red) genes as positive controls, and *dap* gene of *Bacillus subtilis* as negative control in lung (A), heart (B), lymph node (C), kidney (D) and adrenal gland (E) autopsies. Probes against SARS-CoV-2 (green; absent) and *Homo sapiens* *ACE2* (red) were incubated with a lung autopsy collected from a patient diagnosed with influenza A virus subtype H1N1 before COVID-19 outbreak (F). Scale bars represent 20 µm.

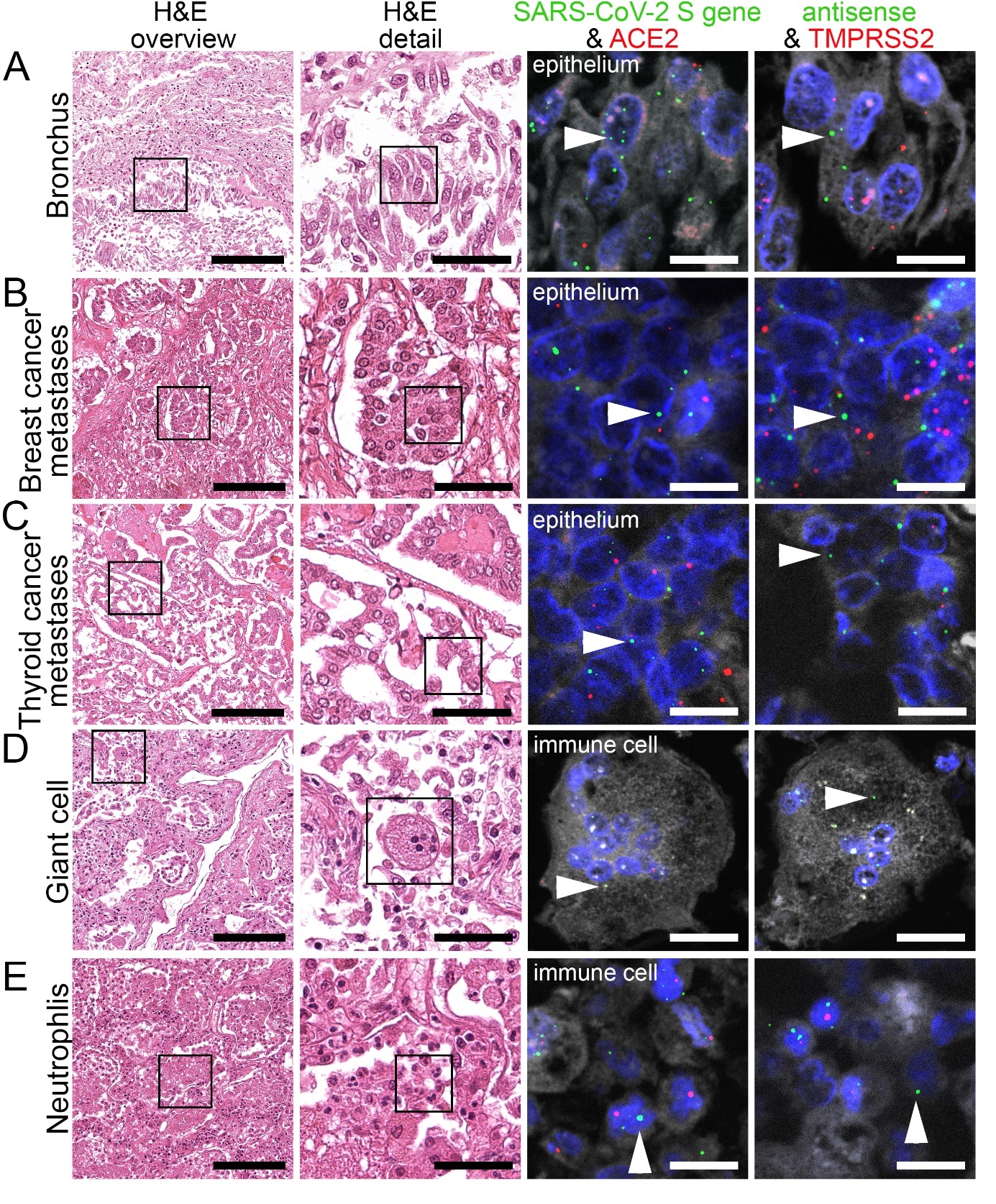

**Supplementary Figure 2. Virus detection in respiratory system by FISH.**

HE stained lung tissue and representative image sections showing FISH co-visualization of RNA sequences either of SARS-CoV-2 S gene genomic RNA (green, arrowhead) and ACE2 (red) or SARS-CoV-2 antisense strand replicating RNA (green, arrowhead) and TMPRSS2 (red). Morphological details are shown in regions of a bronchus (A), metastasis from breast cancer (B) and thyroid cancer (C) and immune cells, i.e., multinucleated giant cells (D) and neutrophils (E). Scale bars represent 200, 50 and 10 µm (A, B, C, D, F) or 20 µm (E), respectively.

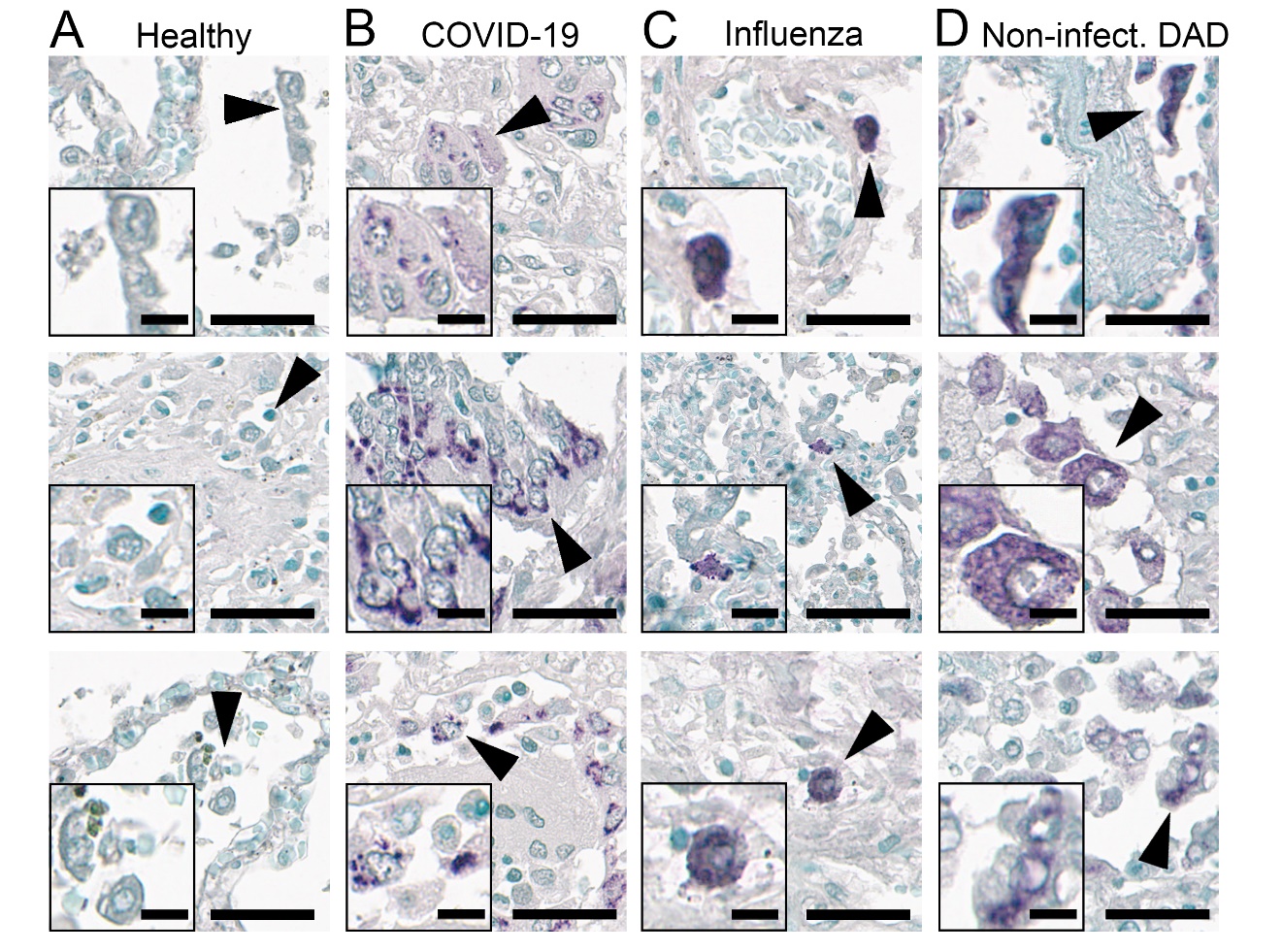

**Supplementary Figure 3. Virus detection in respiratory system by IHC staining.**

Representative IHC staining of SARS spike glycoprotein (#Ab272420) in the autopsy lung tissues collected from different patients in each group of our cohort, consisting of patients without pulmonary disease/ healthy lung (A), COVID-19 patients (B), patients infected with influenza A virus subtype H1N1 (C) or diffuse alveolar damage (non-infectious, D). Scale bars represent 40 µm and 10 µm (insert).

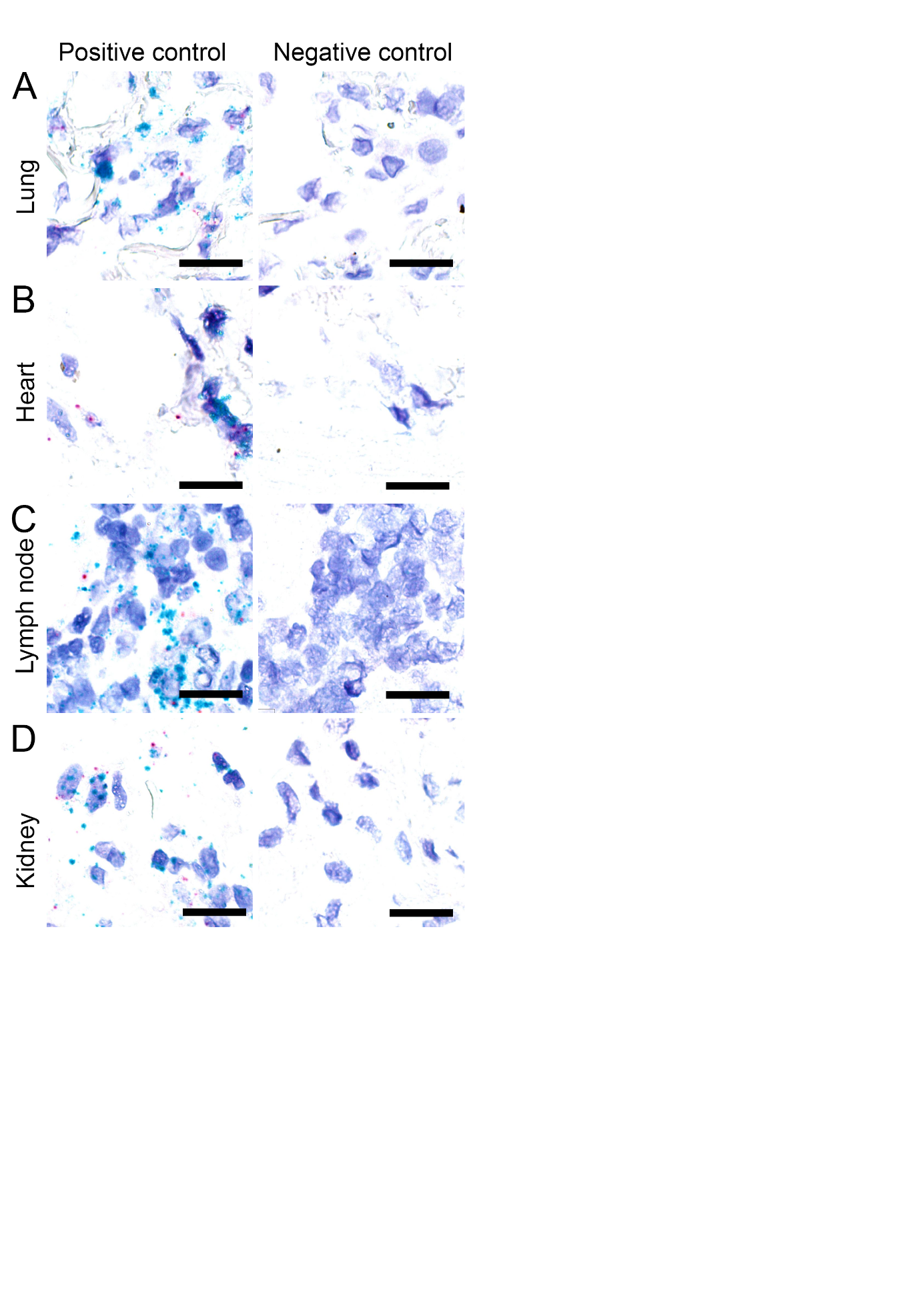

**Supplementary Figure 4. Validation of CISH detection on external human autopsies.**

CISH method was validated by incubation of probes targeting *Homo sapiens* *POLR2A* (cyan) and *PPIB* (red) genes as positive controls, and *dap* gene of *Bacillus subtilis* as negative control in lung (A), heart (B), lymph node (C) and kidney (D) autopsies. Scale bars represent 20 µm.

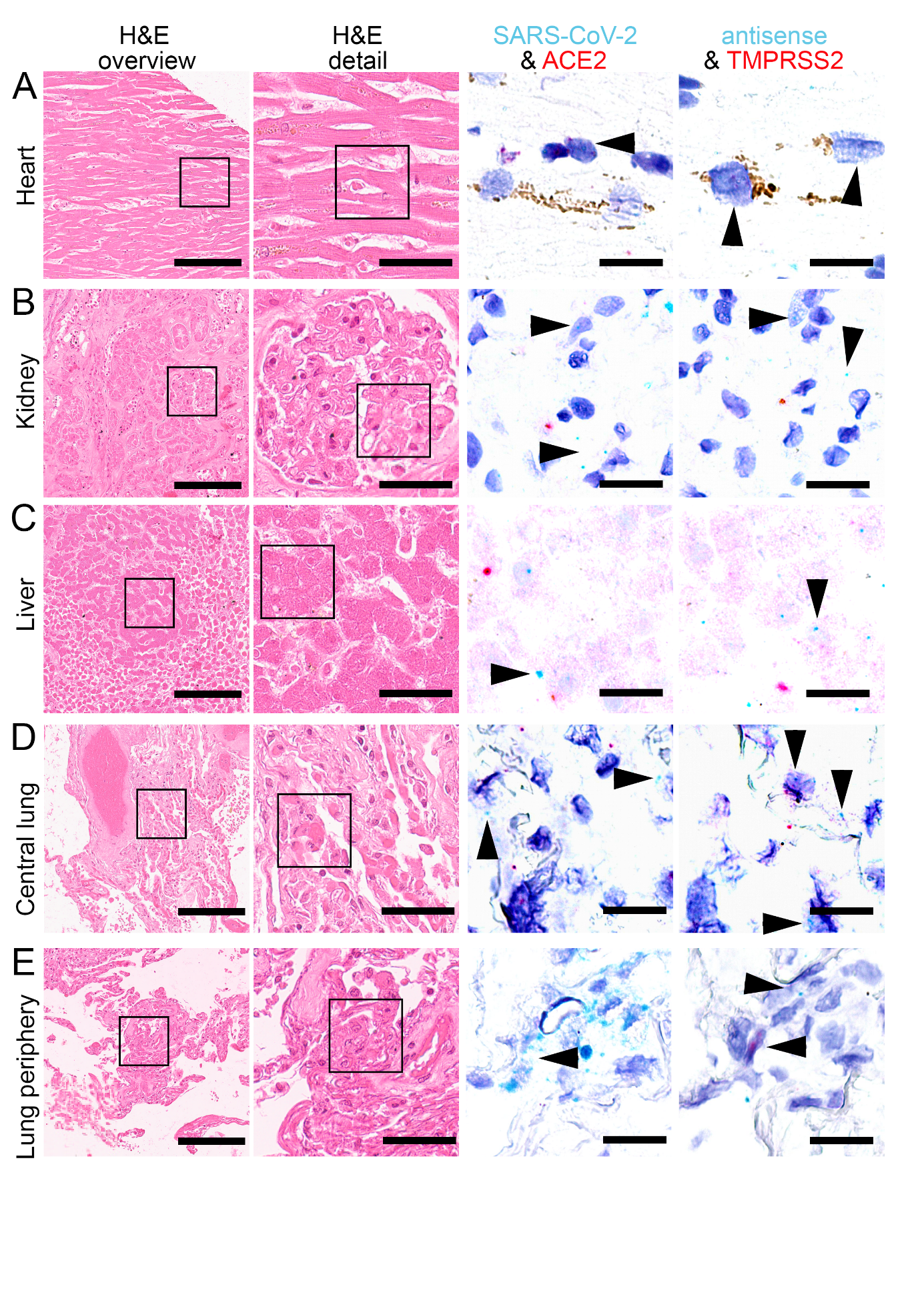

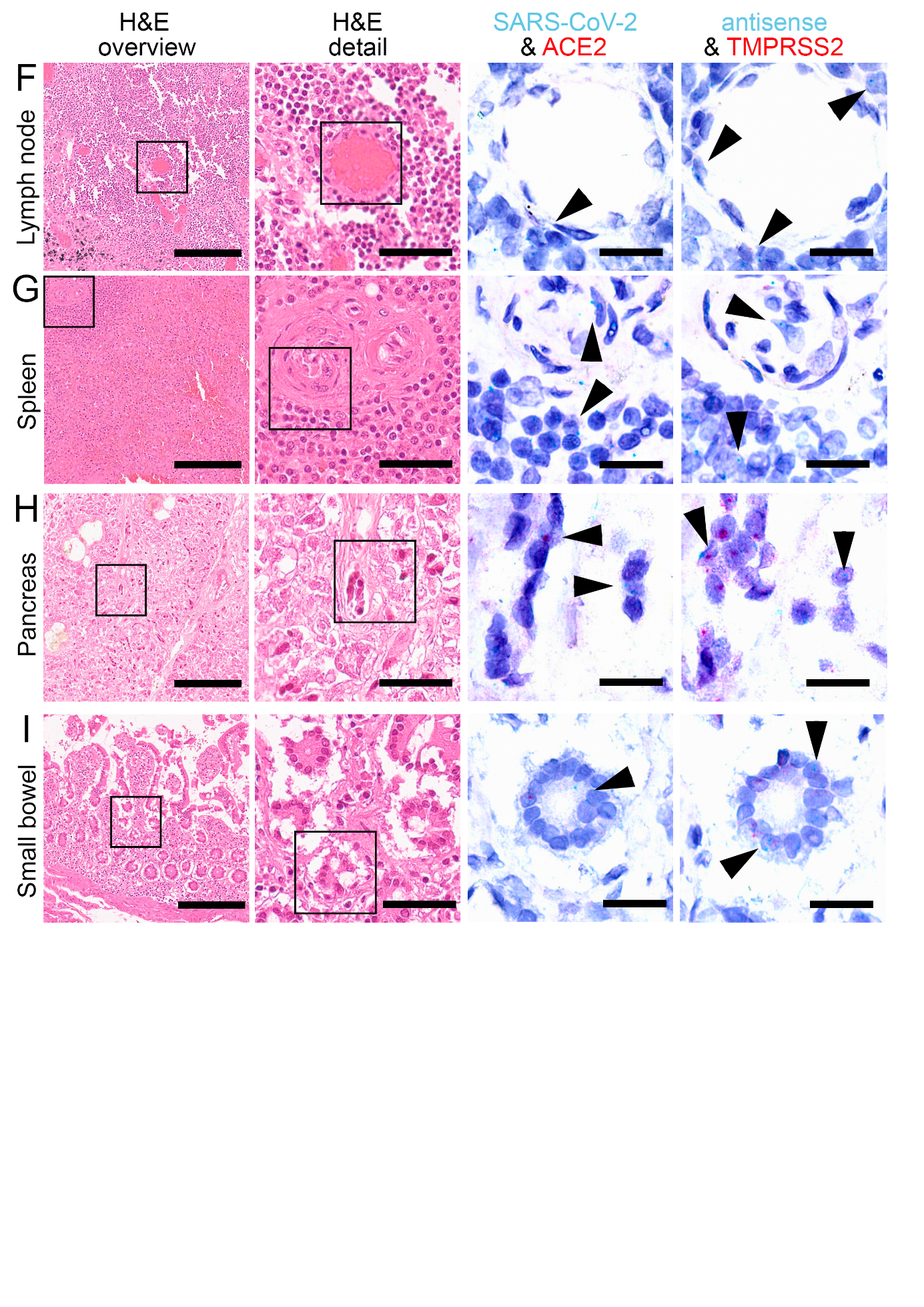

**Supplementary Figure 5. Detection of SARS-CoV-2 sense, antisense, ACE2 and TMPRSS2 on external samples by CISH.**

HE stained tissue and representative image sections showing CISH co-visualization of RNA sequences either of SARS-CoV-2 S gene genomic RNA (cyan, arrowhead) and ACE2 (red) or SARS-CoV-2 antisense strand RNA indicating replicating virus (cyan, arrowhead) and TMPRSS2 (red) in heart (A, cardiomyocyte), kidney (B, glomerulus), liver (C, hepatocyte), central lung (D, alveolus), lung periphery (E, alveolus), lymph node (F, capillary endothelial cells), spleen (G, capillary endothelial cells), pancreas (H, ductal epithelial cells) and small bowel (I, enterocytes). Scale bars represent 200, 50 and 20 µm, respectively.
